# Single nucleotide variants in *Pseudomonas aeruginosa* populations from sputum correlate with baseline lung function and predict disease progression in individuals with cystic fibrosis

**DOI:** 10.1101/2021.10.04.21264421

**Authors:** Morteza M. Saber, Jannik Donner, Inès Levade, Nicole Acosta, Michael D. Parkins, Brian Boyle, Roger Levesque, Dao Nguyen, B. Jesse Shapiro

## Abstract

Complex polymicrobial communities inhabit the lungs of individuals with cystic fibrosis (CF) and contribute to the decline in lung function. However, the severity of lung disease and its progression in CF patients are highly variable and imperfectly predicted by host clinical factors at baseline, CFTR mutations in the host genome, or sputum polymicrobial community variation. The opportunistic pathogen *Pseudomonas aeruginosa* (*Pa*) dominates airway infections in the majority of CF adults. Here we hypothesized that genetic variation within *Pa* populations would be predictive of lung disease severity. To quantify *Pa* genetic variation within whole CF sputum samples, we used deep amplicon sequencing on a newly developed custom Ion AmpliSeq panel of 209 *Pa* genes previously associated with the host pathoadaptation and pathogenesis of CF infection. We trained machine learning models using *Pa* single nucleotide variants (SNVs), clinical and microbiome diversity data to classify lung disease severity at the time of sputum sampling, and to predict future lung function decline over five years in a cohort of 54 adult CF patients with chronic *Pa* infection. The models using *Pa* SNVs alone classified baseline lung disease with good sensitivity and specificity, with an area under the receiver operating characteristic curve (AUROC) of 0.87. While the models were less predictive of future lung function decline, they still achieved an AUROC of 0.74. The addition of clinical data to the models, but not microbiome community data, yielded modest improvements (baseline lung function: AUROC=0.92; lung function decline: AUROC=0.79), highlighting the predictive value of the AmpliSeq data. Together, our work provides a proof-of-principle that *Pa* genetic variation in sputum is strongly associated with baseline lung disease, moderately predicts future lung function decline, and provides insight into the pathobiology of *Pa*’s effect on CF.

**Importance:** Cystic fibrosis (CF) is among the most common, life-limiting inherited disorder, caused by mutations in the CF transmembrane conductance regulator (CFTR) gene. CF causes progressive damage to the lungs, the major cause of morbidity and mortality in CF patients. However, the rate of lung function decline is highly variable across CF patients, and cannot be fully explained using existing biomarkers in the human genome or patient co-morbidities. *Pseudomonas aeruginosa* (*Pa*) is known to evolve and adapt within chronic CF infections. We hypothesized that within-patient *Pa* diversity could affect lung disease severity. In a CF cohort study, we demonstrate the utility of machine learning tools for predictive modeling of baseline lung function and subsequent decline in CF patients using deep within-patient *Pa* amplicon sequencing. Our findings show the potential of these models to identify high-risk CF patients based on *Pa* diversity within the lung.

## Introduction

Cystic fibrosis (CF) is an autosomal recessive disorder caused by mutations in the CF transmembrane conductance regulator (CFTR) gene and is the most common lethal Mendelian disease in populations with European ancestry (Welsh, Ramsey et al. 2001). The resulting lung disease is the major cause of morbidity and mortality in CF patients, with lung failure the most common cause of death (Turcios 2020). However, the rate of disease progression and lung function decline is highly variable across CF populations, and cannot be fully explained by variations in CFTR alleles or other modifier genes (Shanthikumar, Neeland et al. 2019).

While CF airway infections are polymicrobial and microbiome diversity has been associated with lung disease severity in many studies such as (Cox, Allgaier et al. 2010, van der Gast, Flight, Smith et al. 2015, Zhao, Schloss et al. 2012, Coburn, Wang et al. 2015, Cuthbertson, Walker et al. 2020), *Pseudomonas aeruginosa* (*Pa*) is recovered in the majority of adult CF patients and often dominates the CF airway microbiome once established as a chronic infection (Goddard, Staudinger et al. 2012, Zhao, Schloss et al. 2012). Infection with *Pa* in early life is widely recognized to be associated with a greater decline in lung function and mortality (Kosorok, Zeng et al. 2001, Emerson, Rosenfeld et al. 2002, Fothergill, Walshaw et al. 2012). Notably, *Pa* airway infections can persist even with highly effective CFTR-correcting treatment (Hisert, Heltshe et al. 2017, Harris, Wagner et al. 2020).

Over the course of chronic infection in the CF lung, *Pa* undergoes genetic diversification, selection and adaptive evolution, resulting in a genetically and phenotypically diverse population of clonally-related *Pa* within each patient (Tümmler 2006, Smith, Buckley et al. 2006, Bragonzi, Paroni et al. 2009, Mowat, Paterson et al. 2011, Folkesson, Jelsbak et al. 2012, Marvig, Sommer et al. 2015). How this pathoadaptation affects the clinical course of CF lung disease remains poorly understood. We therefore focused on examining the association between *Pa* genetic variation and the severity and progression of lung disease in CF patients with chronic *Pa* infections. We hypothesized that the within-host genetic variation in *Pa* populations during chronic CF lung infections are associated with baseline lung function and subsequent progression (*i*.*e*. decline in lung function), as measured by spirometry.

Within-host mutations can significantly affect the virulence of *Pa* and host responses to *Pa*, (Marvig, Johansen et al. 2013, Marvig, Sommer et al. 2015, Williams, Evans et al. 2015, Klockgether, Cramer et al. 2018, Dettman and Kassen 2021). Previous studies have examined the genetic variation of *Pa* across cohorts of CF patients by performing whole-genome sequencing (WGS) of one or few *Pa* clones isolated from CF sputum samples – an approach that fails to capture the polyclonal nature of *Pa* in the CF lung and is subject to profound sampling bias. While shotgun metagenomic analysis of CF sputum is increasingly used for microbiome analyses (Nelson, Pope et al. 2019, Whelan, Waddell et al. 2020, Lim et al. 2014,), the overwhelming abundance of host derived DNA in samples continues to hamper the ability to resolve within species genetic variation. To overcome these challenges, here we applied a custom-made amplicon sequencing (AmpliSeq) panel of 209 genes in the *Pa* genome previously known to be involved in the pathoadaptation and pathogenesis of CF infections (Supplementary Data 1). The Ion AmpliSeq platform was selected because it provides a means for quantitative and sensitive measurement of single nucleotide variant (SNV) frequencies within the *Pa* population, directly from CF sputum without the need to culture and sequence hundreds of isolates per individual sample.

We then used several machine learning (ML) approaches to classify lung disease severity (at the time of sample collection) and to predict future disease progression (over five years) based on the SNV frequency data from a cohort of 54 adult CF patients with chronic *Pa* infection. ML has been successfully applied to predict phenotypes from genotype data in other model systems (Dias and Torkamani 2019). ML models can explicitly include the interactions and correlations between features (in our case, SNVs), which is particularly common in bacterial population structures in which SNVs are often genetically linked on the same clonal genomic background (Lees, Mai et al. 2020).

Our study provides proof-of-principle evidence that the population of *Pa* in CF sputum includes bacterial genetic biomarkers that are associated with lung disease status and could serve to identify individuals at increased risk of future lung function decline. Additionally, this work identified genetic variation in *Pa* genes that merit further investigation for their potential roles in the pathogenesis of CF lung disease.

## Results

We studied a previously described and well-characterized cohort of young adult CF patients aged 18 to 22 with chronic *Pa* infection (Acosta, Heirali et al. 2018). Using *Pa* SNV frequencies quantified by AmpliSeq in patient sputum, we sought to predict two measures of lung disease severity: (1) baseline lung function (FEVp score) at the time of sputum sample collection, classified as severe or mild, and (2) relative lung function decline in the following five years, classified as rapid or non-rapid. After filtering for AmpliSeq sequencing quality, we excluded 10 patients with low coverage of *Pa*, leaving 54 patients for further analysis (Methods). The clinical and demographic characteristics of the final cohort are summarized in **Table 1**, and the excluded patients were not apparent outliers in their clinical profiles (data not shown). From the filtered sequence data we identified SNVs within the 209 genes represented in the AmpliSeq panel, and estimated the frequency of each SNV within each patient sputum sample. In total across the 54 patient samples, we identified 7,867 synonymous and 4,452 non-synonymous SNVs (Supplementary Data 2). All variants were used for population stratification analysis and only non-synonymous SNVs were used for training ML models.

**Table 1.**
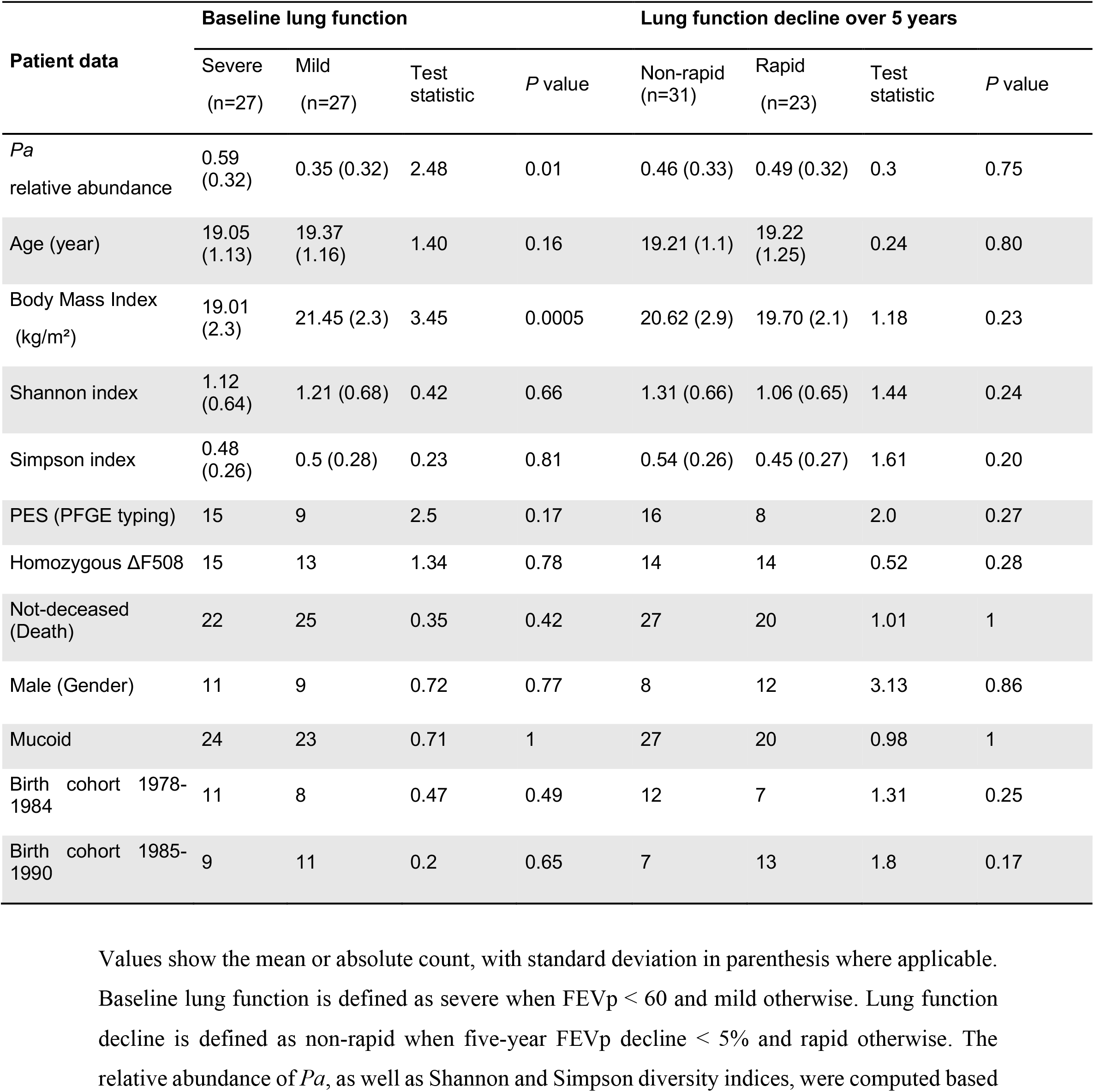
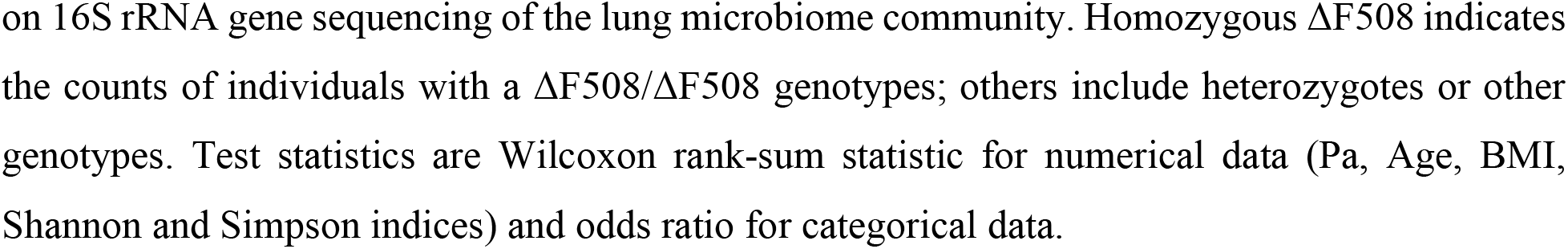
Patient clinical data.

| Patient data | Baseline lung function |  |  |  | Lung function decline over 5 years |  |  |  |
| --- | --- | --- | --- | --- | --- | --- | --- | --- |
|  | Severe<br>(n=27) | Mild<br>(n=27) | Test<br>statistic | *P* value | Non-rapid<br>(n=31) | Rapid<br>(n=23) | Test<br>statistic | *P* value |
| *Pa*<br>relative abundance | 0.59<br>(0.32) | 0.35 (0.32) | 2.48 | 0.01 | 0.46 (0.33) | 0.49 (0.32) | 0.3 | 0.75 |
| Age (year) | 19.05<br>(1.13) | 19.37<br>(1.16) | 1.40 | 0.16 | 19.21 (1.1) | 19.22<br>(1.25) | 0.24 | 0.80 |
| Body Mass Index<br>(kg/m<sup>2</sup>) | 19.01<br>(2.3) | 21.45 (2.3) | 3.45 | 0.0005 | 20.62 (2.9) | 19.70 (2.1) | 1.18 | 0.23 |
| Shannon index | 1.12<br>(0.64) | 1.21 (0.68) | 0.42 | 0.66 | 1.31 (0.66) | 1.06 (0.65) | 1.44 | 0.24 |
| Simpson index | 0.48<br>(0.26) | 0.5 (0.28) | 0.23 | 0.81 | 0.54 (0.26) | 0.45 (0.27) | 1.61 | 0.20 |
| PES (PFGE typing) | 15 | 9 | 2.5 | 0.17 | 16 | 8 | 2.0 | 0.27 |
| Homozygous ΔF508 | 15 | 13 | 1.34 | 0.78 | 14 | 14 | 0.52 | 0.28 |
| Not-deceased<br>(Death) | 22 | 25 | 0.35 | 0.42 | 27 | 20 | 1.01 | 1 |
| Male (Gender) | 11 | 9 | 0.72 | 0.77 | 8 | 12 | 3.13 | 0.86 |
| Mucoid | 24 | 23 | 0.71 | 1 | 27 | 20 | 0.98 | 1 |
| Birth cohort 1978-1984 | 11 | 8 | 0.47 | 0.49 | 12 | 7 | 1.31 | 0.25 |
| Birth cohort 1985-1990 | 9 | 11 | 0.2 | 0.65 | 7 | 13 | 1.8 | 0.17 |

Values show the mean or absolute count, with standard deviation in parenthesis where applicable.
Baseline lung function is defined as severe when FEV<sub>p</sub> < 60 and mild otherwise. Lung function
decline is defined as non-rapid when five-year FEV<sub>p</sub> decline < 5% and rapid otherwise. The
relative abundance of *Pa*, as well as Shannon and Simpson diversity indices, were computed based
on 16S rRNA gene sequencing of the lung microbiome community. Homozygous $\Delta F508$ indicates the counts of individuals with a $\Delta F508/\Delta F508$ genotypes; others include heterozygotes or other genotypes. Test statistics are Wilcoxon rank-sum statistic for numerical data (Pa, Age, BMI, Shannon and Simpson indices) and odds ratio for categorical data.

### Stratification in the *Pa* population

We first quantified the extent of *Pa* population stratification, which can be problematic if there are genetic structures (*e*.*g*. clonally-related clusters or strains) that can be confounded with the lung disease outcomes of interest. If a particular genetic cluster or lineage is associated with worsened lung disease, it then becomes difficult to pinpoint the most likely SNVs associated with the disease outcome because all mutations (whether related to disease or not) in a cluster are correlated. We know *a priori* based on pulsed-field gel electrophoresis (PFGE) typing that our dataset contains a highly prevalent lineage of *Pa* (called <u>P</u>rairie <u>E</u>pidemic <u>S</u>train or PES; sequence type (ST)-192; **Table 1**) suspected to be associated with disproportionate lung disease (Somayaji, Lam et al. 2017). We confirmed this by hierarchical clustering of the *Pa* AmpliSeq data (n=12,319 SNVs, including both synonymous and non-synonymous variants), which revealed two apparent genetic clusters (**Fig. 1a**), one of which was strongly associated with the PES lineage (Fisher exact test, odds ratio=168.0, *P* = 1.1e-09; **Fig. 1b**). The observed *Pa* genetic clusters are also weakly associated with the birth cohort (Chi square test, *P =* 0.0095) which is likely due to unequal prevalence of PES across birth cohorts (Supplementary table S1). No other clinical factor was significantly associated with either genetic cluster (**Fig. 1b**). Importantly, neither cluster is correlated with either baseline lung function (Fisher exact test, p-value: 0.81) or lung function decline (Fisher exact test, p-value: 0.51) (**Fig. 1b**), indicating that these outcomes are unlikely to be confounded by Pa population stratification, and finer-grained predictive modeling is warranted. We also noted that lung disease progression over 5 years (lung function decline) is not significantly correlated with baseline lung function at sample collection (**Fig. 1a**).

**Figure 1.**
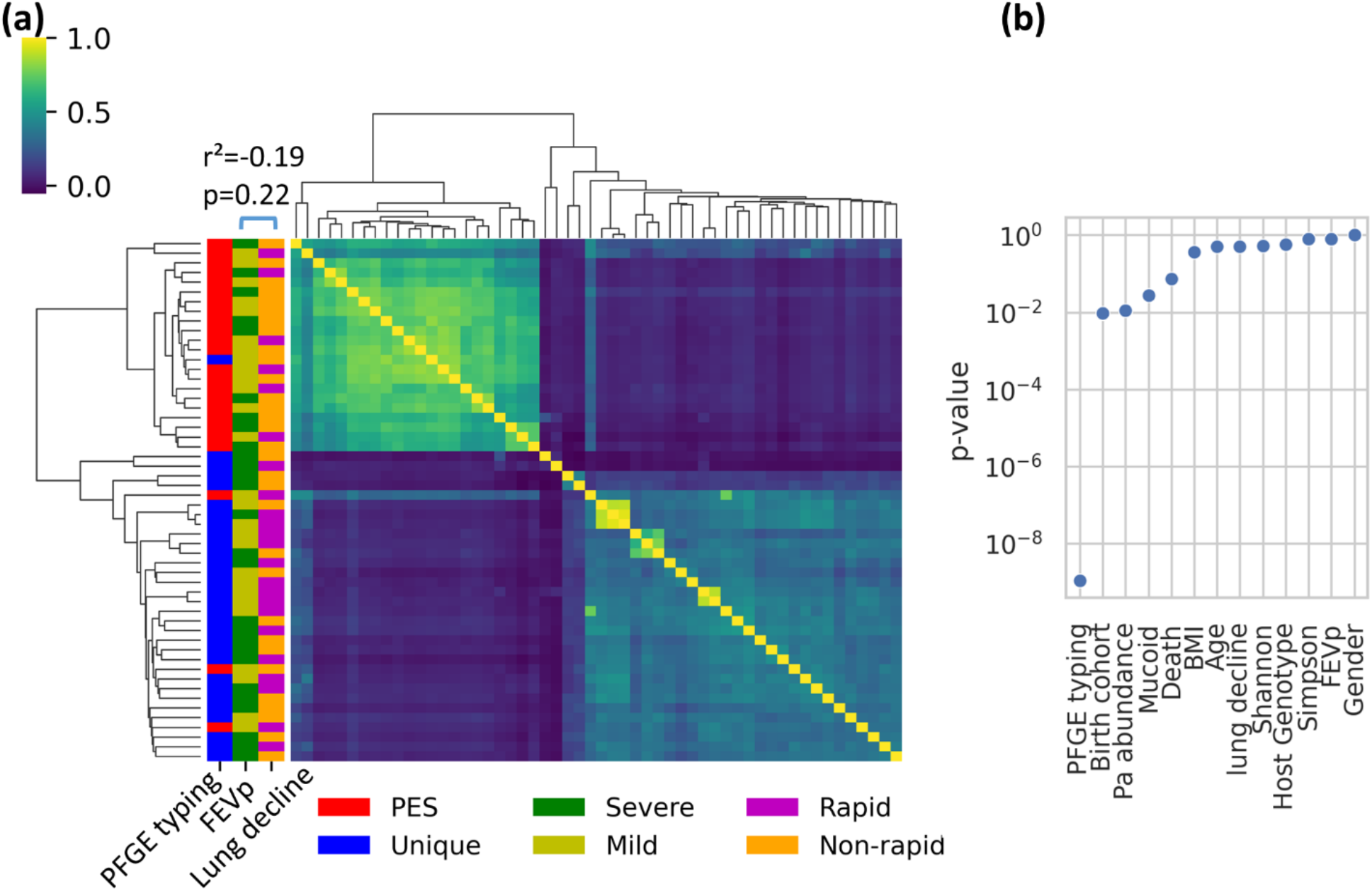
The *Pa* population is stratified into two genetic clusters, neither of which is associated with baseline lung function (FEVp) or lung function decline. (A) Heatmap showing correlations in SNV frequencies between pairs of sputum samples. Strong correlations are yellow; weak correlations in blue. Rows and columns (samples) are ordered by hierarchical clustering. Distribution of baseline lung function measured by FEVp score (27 Severe and 27 Mild individuals), lung function decline (23 Rapid and 31 Non-rapid individuals) and PFGE typing (25 PES and 29 Unique) are presented on the y axis. Baseline lung function and lung function decline over five years are not significantly correlated (Pearson R^2^ score=-0.19, p-value=0.22). (B) *P*-values for the association between clinical data and genetic clusters are determined by *t*-test for numerical data and chi-square test for categorical data (Methods). Only the association between PFGE type (PES or non-PES) is significantly associated with the genetic clusters in panel A (*P* < 0.0045 after Bonferroni correction for multiple tests).

### Genetic and clinical features associated with baseline lung function and lung function decline in CF patients

A common challenge in predicting outcomes from sequence data is the sparsity of the data, that is, relatively few available samples compared to the large number of genetic markers (called “features” in ML context). To address this problem, feature selection has been used to remove non-informative features (*i*.*e*., SNVs and clinical factors) and focus only on the most predictive ones (Mobegi, Cremers et al. 2017, Recker, Laabei et al. 2017, Méric, Mageiros et al. 2018, Macesic, Don’t Walk et al. 2020). We used an ensemble gradient boosting technique for feature selection (Methods). Out of 4,452 non-synonymous SNVs and eleven clinical factors considered, our model selected only 34 SNVs (hereafter called predictor SNVs) and three clinical factors (age, BMI and *Pa* abundance) that account for 99% of the cumulative feature importance (**Fig. 2**). This means that a minimal set of SNVs and clinical factors provides 99% of the information used in predicting baseline lung function at the time of sampling (**Fig. 2a**). An equivalent analysis for lung function decline after five years identified 33 predictor SNVs and the same three clinical factors that contributed to 99% of the cumulative feature importance (**Fig. 2b**). For both baseline lung function and future lung function decline, the phenotype is not simply predicted based on the presence/absence of each SNV, but rather on more subtle information about SNV allele frequencies within patients. In other words, predictive SNVs occur at a range of frequencies, rather than being clustered mainly around 0 or 1 (**Supplementary fig. 1**).

**Figure 2.**
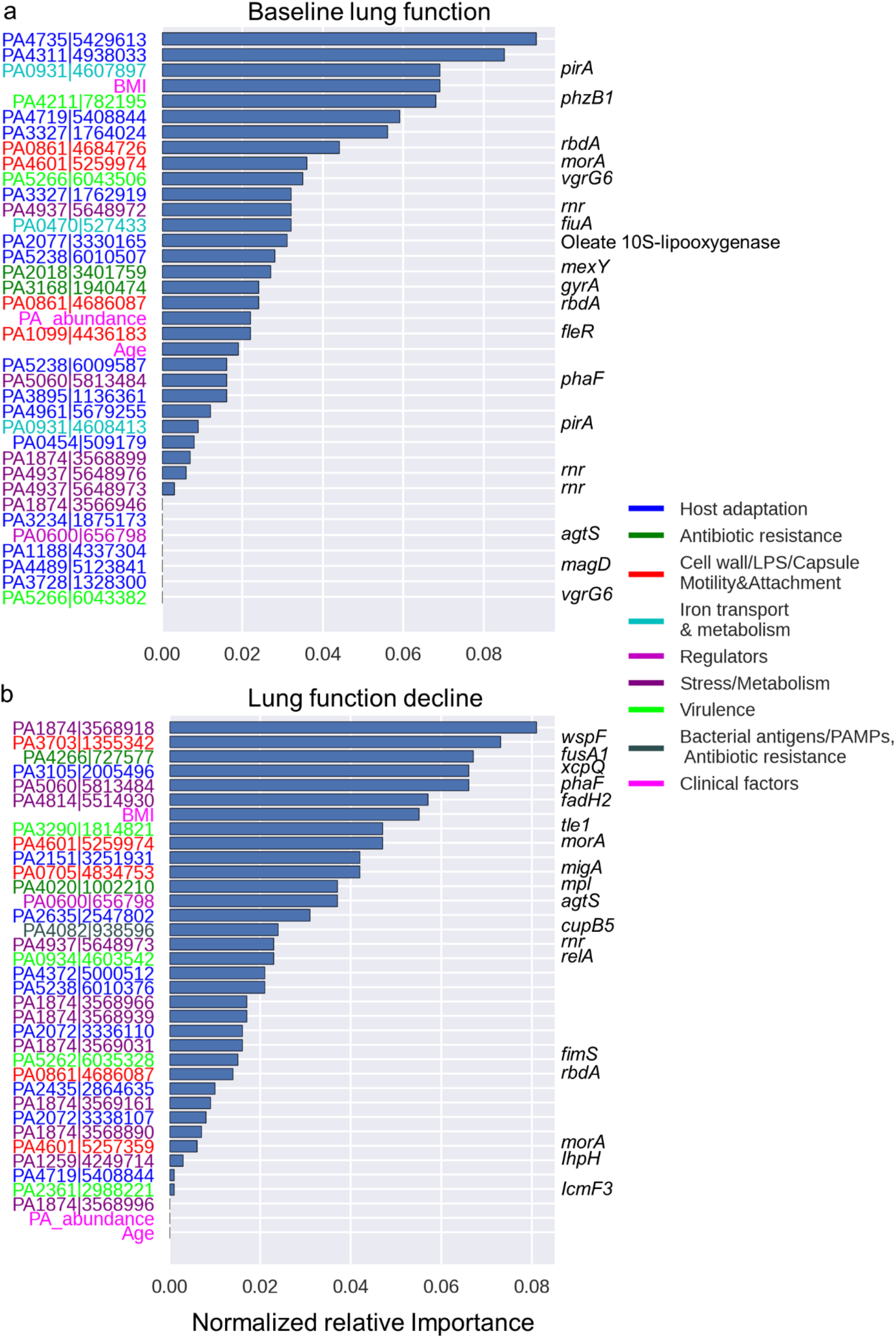
*Pa* genes and clinical factors selected as top predictive features of baseline lung function and lung function decline. Normalized importance of genomic and clinical data that contribute to 99% cumulative relative importance for prediction of (A) baseline lung function at time of sample collection and (B) risk of 5-year progression (lung function decline). On the y-axis, gene identifiers (locus tag|chromosome location based on PES genome) are color-coded based on their functional classification and named genes are shown on the right, when available.

The three selected clinical factors associated with both baseline lung function and lung function decline are body mass index (BMI), *Pa* relative abundance (from 16S rRNA gene amplicon sequence data from a previous study of the same cohort; (Acosta, Heirali et al. 2018), and age (**Fig. 2**). Multiple studies have shown an association between poor lung function and low BMI (Snell, Bennetts et al. 1998, Cystic Fibrosis Foundation 2006, Kumru, Emiralioğlu et al. 2018), high abundance of *Pa* (Cox, Allgaier et al. 2010) and age (Zhao, Hao et al. 2020, Cox, Allgaier et al. 2010). As expected, *Pa* relative abundance also showed a strong negative correlation with Shannon and Simpson microbiome diversity indices (**Supplementary fig. 2**), indicating that *Pa* abundance can be considered as a proxy for lung microbiome diversity in our dataset. However, Shannon and Simpson diversity indices were not selected as predictive features in our model, consistent with a previous work (Acosta et al. 2018; Zhao, Hao et al. 2020). This suggests that, even if low microbiome diversity indices are associated with CF disease progression, the low diversity is likely driven by the dominance of key pathogens such as *Pa*. By identifying previously known clinical determinants of the lung function in CF patients, these results provide validation for the ensemble gradient boosting approach to feature selection.

To interpret the possible roles of *Pa* SNVs in CF lung disease, we classified the known or predicted function of genes containing predictor SNVs (hereafter called predictor genes) into functional categories manually curated based on existing literature. The predictor SNVs with the highest weighted importance for both baseline lung function and future lung function decline outcomes are located within genes that play a role in seven functional categories (**Table 2**). The distribution of predictor genes is generally similar to the distribution of gene functions included in the AmpliSeq panel (**Supplementary fig. 3**). However, the predictor genes for baseline lung function are enriched in iron transport and metabolism (13.4% in baseline lung function predictor genes vs. 1.4% in the AmpliSeq panel, *P =* 0.00018; **Supplementary fig. 3**). The genes encoding the ferric enterobactin receptor (PirA) and the ferrichrome receptor (FiuA) respectively account for 8.9% and 3.7% of the total normalized importance for baseline lung function (**Fig. 2a**), and *pirA* contains multiple predictor SNVs (**Fig. 3a**). In contrast, the predictor genes for lung function decline are enriched in stress/metabolism (33.6 % in lung function decline predictor genes vs. 13.4% in the AmpliSeq panel, *P* = 0.002; **Supplementary fig. 3**). Notably, the hypothetical protein PA1874 accounts for 16.9% of the total normalized importance for lung function decline prediction and includes 7 out of 33 predictor SNVs (**Fig. 2b, Fig. 3b**), as well as two predictor SNVs for baseline lung function (**Fig. 3a**). This hypothetical protein has also been shown to play a role in resistance of *Pa* to multiple antibiotics (Zhang and Mah 2008). The PA4937 gene, which encodes an RNase R exoribonuclease involved in stress/metabolism, is also a predictor of both baseline lung function and subsequent lung function decline, with multiple predictor SNVs (**Fig. 2a, Fig. 3a**). The two genes PA0861 (*rbda*) and PA4601 (*morA*), which encode regulators of genes involved in the bacterial cell wall, LPS, capsule, motility and attachment, are also among the important predictor genes for both baseline lung function and lung function decline (**Fig. 2**).

**Table 2.** Functional classification of predictor genes used for prediction of baseline lung function and lung function decline.

|  | Baseline lung function<br>predictor genes | Lung function decline<br>predictor genes | Shared genes |
| --- | --- | --- | --- |
| **Host adaptation** | PA0454\|*conserved hypothetical protein*<br>PA1188\|*hypothetical protein*<br>PA2077\|*oleate 10S-lipoxygenase*<br>PA3234\|*probable sodium:solute symporter*<br>PA3327\|*probable non-ribosomal peptide synthetase*<br>PA3728\|*hypothetical protein*<br>PA3895\|*probable transcriptional regulator*<br>PA4311\|*conserved hypothetical protein*<br>PA4489\|*magD*<br>PA4735\|*hypothetical protein*<br>PA4961\|*hypothetical protein* | PA2072\|*conserved hypothetical protein*<br>PA2151\|*conserved hypothetical protein*<br>PA2435\|*probable cation-transporting P-type ATPase*<br>PA2635\|*hypothetical protein*<br>PA3105\|*xcpQ*<br>PA4372\|*hypothetical protein* | PA4719\|*probable transporter*<br>PA5238\|*probable O-antigen acetylase* |
| **Antibiotic resistance** | PA2018\|*mexY*<br>PA3168\|*gyrA* | PA4020\|*mpl*<br>PA4082\|*cupB5*<br>PA4266\|*fusA1* |  |
| **Cell wall, LPS, capsule, motility & attachment** | PA1099\|*fleR* | PA0705\|*migA*<br>PA3703\|*wspF*<br>PA4082\|*cupB5* | PA0861\|*rbdA*<br>PA4601\|*morA* |
| **Iron transport and metabolism** | PA0470\|*fiuA*<br>PA0931\|*pirA* |  |  |
| **Regulators** |  |  | PA0600\|*agtS* |
| **Stress/ metabolism** |  | PA1259\|*lhpH*<br>PA4814\|*fadH2* | PA1874\|*hypothetical protein*<br>PA4937\|*rnr*<br>PA5060\|*phaF* |
| **Virulence** | PA4211\|*phzB1*<br>PA5266\|*vgrG6* | PA0934\|*relA*<br>PA2361\|*icmF3*<br>PA3290\|*tle1*<br>PA5262\|*fimS* |  |

**Figure 3.**
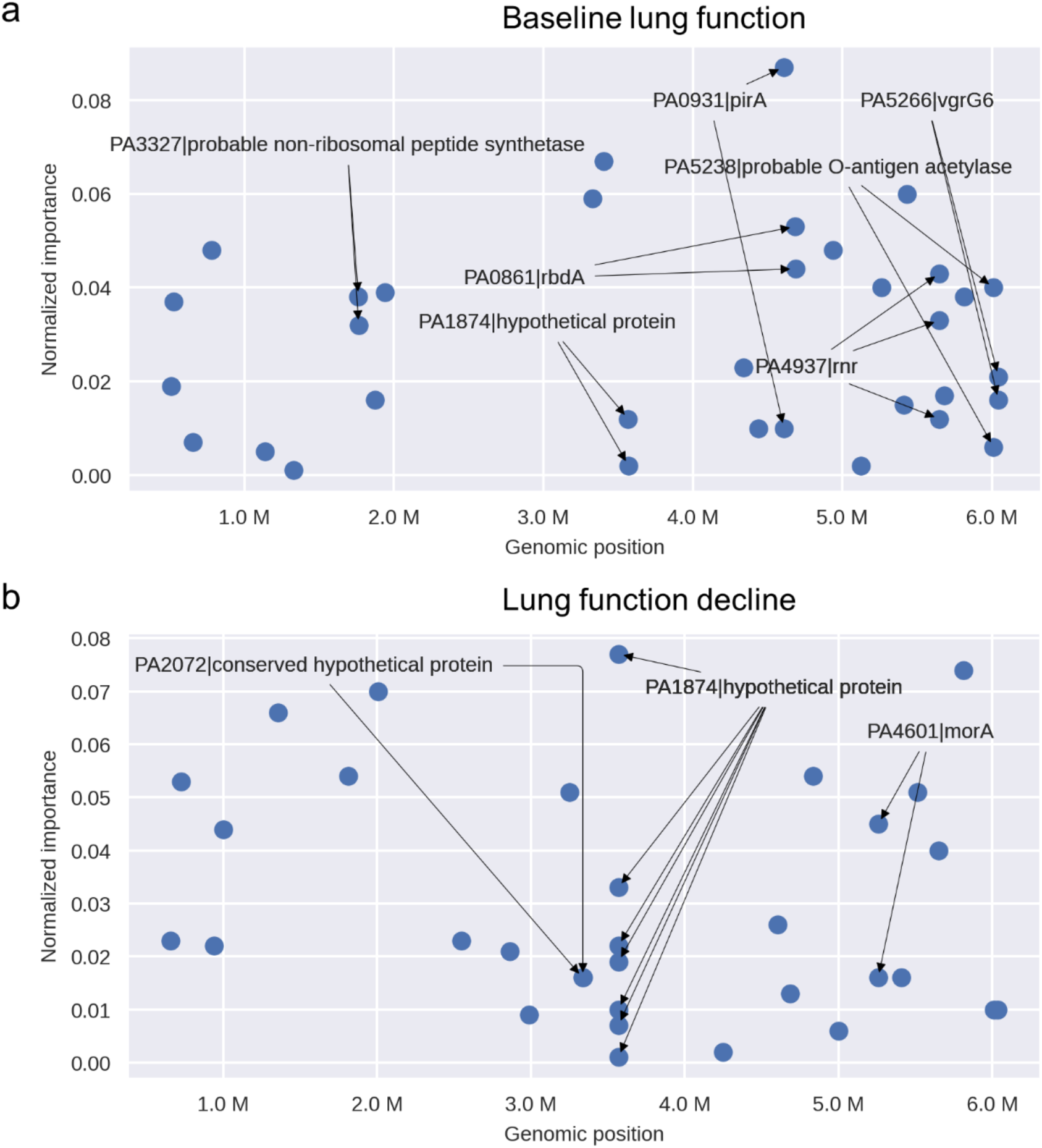
Genomic locations and importance of genes containing predictor SNVs. (A) Genes containing SNVs predictive of baseline lung function. (B) Genes containing SNVs predictive of lung function decline. Genes including multiple SNVs are shown with arrows.

### Predicting lung disease severity and progression in individuals with CF using genetic and clinical factors

Having identified *Pa* SNVs predictive of disease outcomes, we evaluated their predictive performance using the area under receiver operating characteristic (AUROC) curve and other standard metrics (Methods). A genetic programming algorithm identified logistic regression as the best predictive model for both baseline lung function and lung function decline. To confirm this result, we compared the performance of logistic regression with three other common ML algorithms including support vector machines (SVM), random forests, and extreme gradient boosting (XGBoost). It should be noted that while we used an ensemble LGBM model (Methods) for feature selection, we did not use it for predictive modeling to avoid overfitting (*i*.*e*. to confirm that our feature selection method is not biased toward selecting features that could be used only by one specific ML model). Logistic regression had the best predictive performance for both phenotypes (**Table 3, Fig. 4**). Specifically, cross-validation (Methods) showed that logistic regression was the most accurate and precise model for both baseline lung function, with an average AUROC score of 0.87 (95% CI, 0.84-0.90), and for lung function decline, with a score of 0.74 (95% CI: 0.71-0.78; **Table 3, Supplementary table 2**). Linear logistic regression is a simple classification model that makes it reasonably robust against overfitting (Kuhn and Johnson 2013). The second-best method was SVM, also a linear model (**Supplementary table 2**). Across all models, the baseline lung function phenotype was more accurately predicted than lung function decline, consistent with predictions becoming more uncertain further into the future (**Table 3, Supplementary table 2**). Importantly, all models could predict both phenotypes significantly better than expected by chance (compared to a permutation test using data with shuffled outcome labels; **Fig. 4a, c, Supplementary fig. 4**).

**Table 3.** Performance of logistic regression in predicting baseline lung function and lung function decline using genomic data only, or a combination of genomic and clinical data. See Methods for descriptions of the performance metrics.

|  |  | Genomic data<br>(95% CI) | Genomic and clinical<br>data (95% CI) |
| --- | --- | --- | --- |
| **Baseline**<br>**lung**<br>**function** | AUROC | 0.87 (0.84, 0.9) | 0.92 (0.84, 1.00) |
|  | bACC | 0.81 (0.78, 0.84) | 0.83 (0.72, 0.94) |
|  | Accuracy | 0.81 (0.78, 0.84) | 0.83 (0.72, 0.94) |
|  | F1 | 0.81 (0.78, 0.83) | 0.83 (0.72, 0.94) |
|  | Precision | 0.83 (0.81, 0.86) | 0.84 (0.73, 0.94) |
|  | Recall | 0.81 (0.78, 0.84) | 0.83 (0.72, 0.94) |
| **Lung**<br>**function**<br>**decline** | AUROC | 0.74 (0.71, 0.78) | 0.79 (0.70, 0.88) |
|  | bACC | 0.63 (0.59, 0.66) | 0.66 (0.59, 0.74) |
|  | Accuracy | 0.64 (0.60, 0.67) | 0.67 (0.60, 0.75) |
|  | F1 | 0.62 (0.58, 0.65) | 0.66 (0.58, 0.74) |
|  | Precision | 0.65 (0.61, 0.69) | 0.69 (0.60, 0.78) |
|  | Recall | 0.64 (0.60, 0.67) | 0.67 (0.60, 0.75) |

**Figure 4.**
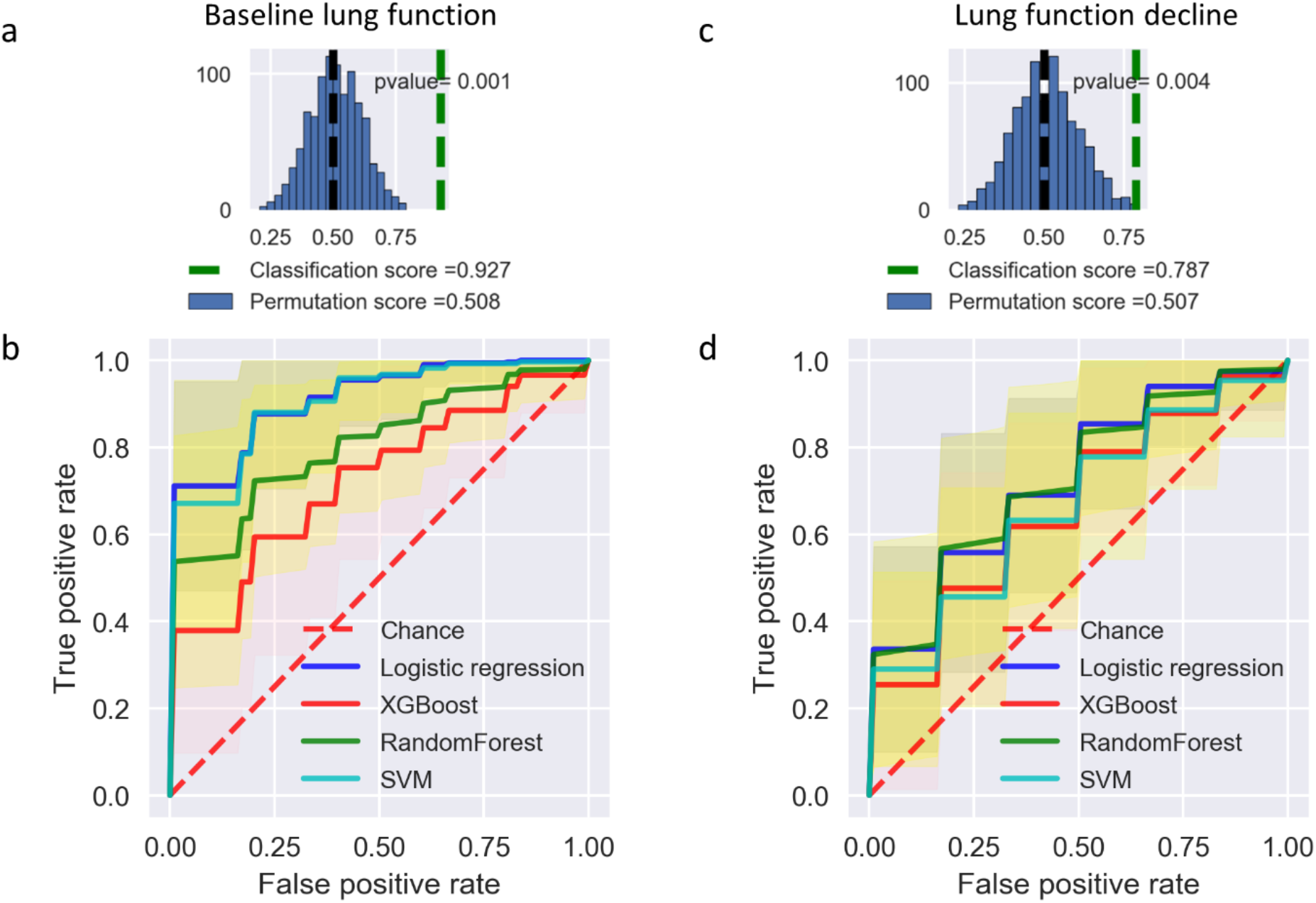
Predictive models of baseline lung function and lung function decline perform significantly better than expected by chance. (A) Classification score of baseline lung function using logistic regression (green dashed line) is significantly higher than expected based on permuted data (mean shown in black dashed line). (B) Average AUROC scores of different ML models to predict baseline lung function, compared to chance (permuted sample labels). Shading indicates the 95% confidence interval. (C) Classification score of lung function decline using logistic regression compared to permuted data. (D) Average AUROC scores for lung function decline prediction.

Clinical factors have been traditionally used to predict lung disease progression in CF patients (Alaa and van der Schaar 2018). We therefore assessed if integrating clinical factors could improve upon the predictions based on *Pa* AmpliSeq data alone. The three clinical factors (BMI, *Pa* abundance and age) identified by our feature selection approach are previously recognized as factors affecting lung function in CF patients. Including these three clinical factors in our predictive models led to modest (∼5% increase in AUROC) performance increases for both baseline lung function and lung function decline outcomes across the four ML models (**Table 4, Supplementary table 2**). We conclude that, while these clinical factors are useful, most of the predictive power comes from the *Pa* genetic data.

Lack of generalizability is one of the main limiting factors for the translation of prediction models into clinically useful diagnostics. Machine learning models often have low generalizability (i.*e*. “overfit”) in scenarios where the model performs well on the dataset used to train the model, but fails to achieve similar prediction accuracy on new data. We plotted learning curves to assess how logistic regression predictions improved by training on more data (Raschka 2018). We found that the performance difference between training and testing data decreases as sample size increases (**Supplementary Figure 5**). There were no major differences in prediction accuracy of training and testing datasets (**Supplementary Figure 5**) which suggests the model does not suffer from significant overfitting. We also noted that cross validation scores for both baseline lung function and lung function decline models continued to increase for the testing dataset as more data was used for model training (**Supplementary Figure 5**) which suggests the model could be further improved with more data.

## Discussion

Considering the critical role of *Pa* in CF-related morbidity and mortality, here we established a link between *Pa* genetic diversity and lung disease severity in a cohort of CF young adults with chronic *Pa* infections. Despite a modest sample size, our study provides a proof of principle demonstrating the utility of ML models for predictive modeling of lung function severity and decline in CF patients using bacterial genetic and clinical data. Although our models do not appear to be severely overfit, fully validating their predictive performance will require independent cohorts. We also identified potential genetic biomarkers associated with lung disease severity. Overall, our findings provide evidence that ML models can identify CF individuals at high-risk for poor *Pa* infection outcomes using *Pa* genetic data.

Our work is based on a subset of samples from a previously described cohort study that identified dominance of *Pa* in the sputum microbiome (and the resulting reduction of community diversity) as a predictor of lung function decline in a cohort of young CF adults (Acosta, Heirali et al. 2018). Here we focused on a subset of patients with a lung microbiome dominated by *Pa*. While these patients are already at increased risk of lung disease, we found that the severity of disease at the time of sampling and five years into the future could be predicted based on genetic variation within the infecting *Pa* population. Even within this patient cohort in which *Pa* was present, we replicated the finding that *Pa* relative abundance is associated with disease severity and progression – although it is a less important biomarker than many SNVs within the *Pa* genome. This suggests that genetic variation in dominant pathogens can significantly complement and improve upon predictions of disease status based on the microbiome. Along these lines, another recent study showed that the *Pa* genomic data can predict pathogenicity in mouse models (Pincus, Ozer et al. 2020).

We recognize that, in addition to variation in the host genome, the polymicrobial community inhabiting the CF lung has been identified as an important modifier of disease progression. Numerous studies of the lung microbiome have shown an association between decreasing microbial community diversity and worsening lung function (Cox, Allgaier et al. 2010, van der Gast, Flight, Smith et al. 2015, Zhao, Schloss et al. 2012, Coburn, Wang et al. 2015, Cuthbertson, Walker et al. 2020), as well as progression to end-stage lung disease (Acosta, Heirali et al. 2018). However, microbiome diversity may have limited predictive value as there is high interpersonal variability in lung microbiomes (Cuthbertson, Walker et al. 2020), and a large number of adult CF individuals have microbiomes dominated by pathogens such as *Pa*. Zhao, Hao et al. (2020) also recently showed that microbiome composition data does not improve machine learning (ML) prediction performance compared with using only clinical factors. Our results suggest that genetic diversity within key pathogens like *Pa* could complement or even supersede microbiome community diversity for predicting clinical outcomes in specific patient subsets.

Limitations of our study include a relatively small sample size of patients (*N=*54) from a single cohort, and a relatively small number of targeted genes (*N=*209) included in the AmpliSeq panel. As such, we consider our work a proof of concept that could be improved upon in larger cohorts and by including more loci in the *Pa* genome. Indeed, learning curves showed that predictive accuracy is likely to improve with more samples. Although we performed cross-validation by subsampling our 54 patients for model training and testing, the model should ideally be tested on a completely independent cohort to assess its real-world predictive value. Despite these limitations, the models made significantly better predictions than expected by chance. As expected, predicting lung function decline five years into the future proved more challenging than doing so at the time of sampling. Still, our results provide a key first step toward clinical diagnostics of patients most at risk of lung function decline.

As with any genotype-phenotype association method, our approach does not fully guarantee causal relationships, and rather provides candidate genes for further experimental testing. Our study is further complicated by the fact that the phenotypes of interest (i.e., baseline lung function and future lung function decline) are complex host phenotypes, while the genotype data comes from only *Pa*. It is therefore unclear to what extent *Pa* SNVs play a causal role in lung function decline, or merely serve as useful biomarkers. Regardless, we were able to pinpoint SNVs in several genes of interest. This was feasible because the strong population stratification of *Pa* into PES and non-PES lineages was fortunately not associated with the disease outcomes of interest. This allowed us to identify SNVs in several genes that provided independent biomarkers of disease.

Several candidate genes containing SNVs predictive of disease status and progression were identified as targets for further investigation. We note that genes in the AmpliSeq panel were selected *a priori* for their known involvement in virulence, disease progression, or within-patient evolution. However, it was not known *a priori* which, if any, of these genes would contains SNVs predictive of lung function or decline. For example, we found that baseline lung function predictor SNVs are enriched in genes involved in iron transport and metabolism. The AmpliSeq panel only included three iron-related genes, of which two (*pirA* and *fiuA*) contained SNVs associated with baseline lung function. Updated AmpliSeq panels or whole-genome sequencing, along with targeted experimental studies, could be used to test the hypothesis that variation in these genes plays a role in disease progression. Multiple studies have shown competition for iron to be key for the survival and virulence of many of the pathogens that reside in the CF lung, including *Pa* (Bouvier 2016, Firoz, Haris et al. 2021). We also found that SNVs predictive of lung function decline are enriched in genes involved in stress/metabolism. Notably, the gene PA1874 includes seven predictor SNVs comprising 16.9% of total feature importance for lung function decline, and two predictor SNVs for baseline lung function prediction, suggesting its general importance in disease severity and progression in CF patients. PA1874 encodes a multidrug efflux pump involved in biofilm-dependent resistance to antibiotics including tobramycin, gentamicin, and ciprofloxacin (Zhang and Mah 2008; Poudyal and Sauer 2018), and could be a potentially promising biomarker of CF disease severity, which merits further investigation.

Among the set of clinical factors studied, BMI, *Pa* abundance, and age were identified as important predictors of both baseline lung function and lung function decline. These are all known risk factors for CF disease severity and progression (Acosta et al. 2018, Snell, Bennetts et al. 1998, Kumru, Emiralioğlu et al. 2018, Cox, Allgaier et al. 2010, Zhao, Hao et al. 2020). By including these features in our prediction models, we noted a moderate increase across all the measured metrics relative to using only AmpliSeq data. These results are in line with previous studies showing the improvement of ML-based phenotype prediction by adding relevant clinical data (MacFadden, Melano et al. 2019, Pincus, Ozer et al. 2020). We note that clinical factors only modestly improved the performance of the models (∼5%), highlighting the rich information and predictive value of the *Pa* AmpliSeq data alone.

In summary, our study demonstrates that SNVs in the *Pa* genome, assayed with an AmliSeq panel and identified by ML models, can be powerful predictors of lung disease severity and progression in CF patients with chronic *Pa* infections. Even though this disease outcome is affected by multiple microbial, host genetic and environmental factors, *Pa* SNVs add complementary predictive value. With additional genetic and clinical data, our ML model could be further fine-tuned and eventually used as a biomarker to preemptively identify individuals with CF at high-risk for more aggressive observation and treatment.

## Materials and methods

### Patient selection, sample and clinical data collection

The Calgary biobank includes frozen whole sputum samples prospectively collected from individuals with CF followed at the Calgary Adult CF clinic from 1998 to 2017, as described previously (Acosta, Heirali et al. 2018, Acosta, Whelan et al. 2017). A cohort of 104 individuals between the ages of 18 and 22 with sputum available from the Calgary biobank was previously characterized (Acosta, Heirali et al. 2018). For this study, we selected from this cohort all individuals with sputum cultures positive for *Pa* (64 out of 104 patients). Out of these 64 samples, 54 yielded AmpliSeq data of sufficient depth (>10X average depth of coverage of the targeted genes) and were retained for further analysis. Clinical data collected for each patient is outlined in **Tables 1** and **S1** and includes age, gender, body mass index, CFTR genotype, birth cohorts, and microbiology (mucoid phenotype, *Pa* relative abundance, microbiome diversity indices). The study was carried out with the approval from the Research Ethics Boards from the University of Calgary (15-0854) and McGill University Health Centre (15-623).

As a measure of lung disease severity at the time of sputum collection, we used the spirometric measure of forced expiratory volume in one second, percent predicted (hereafter referred to as ‘baseline lung function’ and noted FEVp) a standard measure of lung function normalized for age, height, and self-identified gender and ethnicity. Baseline lung function was categorized as severe for FEVp < 60%, and mild for FEVp ≥ 60%. Long-term lung function decline (hereafter noted as ‘lung function decline’) was measured using the relative rate of FEVp decline (determined by subject-specific constructed linear regressions over the 5 years following sputum collection as described in Acosta, Heirali et al. (2018). Lung function decline was categorized as ‘rapid’ when the 5-year FEVp decline was >5%, and ‘non-rapid’ when less than or equal to 5%.

### Sputum DNA extraction and microbiome analyses

Genomic DNA was extracted from a single biobanked sputum sample per patient as previously described (Acosta, Heirali et al. 2018), and used as template for 16S rRNA gene amplicon and Ion AmpliSeq sequencing. The Prairie Epidemic Strain (PES) genotype, a highly prevalent strain in our study population, was identified by pulse field gel electrophoresis (PFGE) and/or multi-locus sequence typing (MLST) (Parkins, Glezerson et al. 2014). For microbiome analysis, bacterial communities in CF sputum and reagent blanks were characterized by amplification and sequencing the V3-V4 region of the 16S rRNA gene, as previously described (Acosta, Heirali et al. 2018). The sequencing reads were then processed to identify operational taxonomic units (OTUs) (Acosta, Whelan et al. 2017). Relative *Pa* abundance was determined as the proportion of *Pseudomonas* reads relative to the total 16S reads.

### Ion AmpliSeq panel design and sequencing

The AmpliSeq panel targeted 209 *Pa* genes previously implicated in pathogenicity, antimicrobial resistance and within-host pathoadaptation during chronic infection (Supplementary Data 1). The AmpliSeq primer panel (generated by Life Technologies, Carlsbad, CA, U.S.A.) was designed by the AmpliSeq Custom Services (White Glove, Thermo Fisher Scientific) to provide high sequencing coverage of the target genes based on the *Pa* PAO1 genome (NCBI accession number: GCA_000006765.1), with 100% breadth of coverage for 205 genes and >96% in 4 genes, based on the tiling of amplicons. Four additional genome assemblies of *Pa* clinical isolates (GCF_004375495.1, GCF_004374685.1, GCF_004374275.1 and the PES genome (NCBI BioProject: PRJNA750451) were also evaluated along with PAO1 for the optimization of primer design, tiling and pooling to achieve maximal target coverage by the primer panel with minimal misalignments and homology with the human genome.

AmpliSeq libraries were constructed using the Ion AmpliSeq™ Library kit 2.0 and IonCode™ barcode set with the following modifications. SparQ magnetic beads (Quantabio) were used for purification, and individual libraries were quantified using the Quant-iT™ PicoGreen™ dsDNA Assay Kit (ThermoFisher). Samples were mixed in equimolar proportions and the pooled library (200 pM) was loaded on an Ion Chef for template preparation using HiQ reagents. The P1 v3 chips were sequenced using an Ion Proton sequencer (500 flows) with P1 HiQ sequencing reagents following manufacturer’s instructions.

### AmpliSeq variant calling

The quality of AmpliSeq sequencing was confirmed using TorrentSuite™ software (v.5.2, Thermo Fisher Scientific Inc.). Raw sequencing reads were trimmed based on per-base phred quality score cutoff (‘q’ flag) of 18, window size of 1 base pair and minimum remaining sequence length (‘l’ flag) of 19 using fastq-mcf (v.1.04.636) (Aronesty 2013). Reads were aligned to the PES genome (CP080405) using BWA MEM (Li, 2013), and the alignments were sorted and indexed using SAMtools (v.1.9) (Li, Handsaker et al. 2009). Samples with average sequencing depth <= 10X across the target genes were discarded, leaving 54 samples for further analysis. Single-nucleotide variants (SNVs) with minimum mapping quality of 20, minimum base quality of 18 and minimum coverage of 10x were then identified using VarScan 2 (Koboldt, Zhang et al. 2012) and functional consequences of each SNV were inferred using snpEFF (v.2.4.2) (Cingolani, Platts et al. 2012). The SNV allele frequencies (ranging from 0 to 1) at each polymorphic site covered by the AmpliSeq panel were used to generate a SNV frequency matrix, with samples as rows and nucleotide positions as columns. For baseline lung function (measured based on FEVp score) and lung function decline (disease progression) prediction analysis, all synonymous variants were filtered out and only non-synonymous variants (including nonsense and missense mutations, frameshift deletions and insertions) were used (Supplementary Data 2). All SNVs (including synonymous sites) were included for population stratification analyses.

### Bacterial population stratification

Population stratification in *Pa* was evaluated by calculating pairwise Pearson correlation coefficients between sputum samples based on the SNV frequency matrix followed by determination of distinct genome subgroups using hierarchical agglomerative clustering implemented in SciPy (Virtanen, Gommers et al. 2020) and visualized using the python seaborn package (Waskom, Botvinnik et al. 2017). This identified two major subclusters of *Pa*, one of which was significantly enriched in PES strains. To determine if any clinical factors were associated with these subclusters, we used t-tests for continuous variables including age, body mass index (BMI), Shannon and Simpson diversity indices and *Pa* abundance in the sputum sample. For binary variables including PFGE typing (PES or not), gender, host CFTR genotype, death, mucoid presence/absence status, baseline lung function and lung disease progression (lung function decline), we used a Fisher Exact Test. A Chi-square test was used for the multi-categorical birth cohort factor.

### Feature selection

In a machine learning context, a feature is defined as an individual measurable characteristic of an observed phenomenon. In this study, the features considered are *Pa* genetic variants (SNV frequencies) identified by the AmpliSeq panel and the clinical factors linked to the study patients (Supplementary table 1). In order to reduce the high-dimensionality of the dataset (i.e. high ratio of features to sample size), a feature selection approach was applied using feature-selector v1.0.0 (Koehrsen 2019). Briefly, 50 rounds of a gradient boosting ensemble method implemented in LightGBM (Ke, Meng et al. 2017) were conducted on the training dataset sampled by a bootstrap approach (43 samples in training set and 11 in test set in each bootstrap). The feature importance (i.e. scores assigned to each input feature indicating the relative importance of the feature when making a prediction) were averaged over the 50 bootstraps. The set of features required to obtain 99% cumulative relative importance were kept to perform prediction analysis and the remaining features were discarded. The enrichment of predictor SNVs across functional gene categories relative to the total genes in the AmpliSeq panel were measured using a Fisher exact test with a family-wise error rate of 0.05 adjusted for multiple testing using Bonferroni method.

### Training predictive model of lung function

The selected SNVs and clinical features were then used to train independent prediction models for baseline lung function and disease progression (lung function decline) in the CF patient cohort. To identify the best prediction model, a genetic programming algorithm implemented in TPOT (Le, Fu et al. 2019) was used. TPOT attempts to identify the machine learning prediction pipeline with the best performance (i.e. cross-validation using the area under the receiver operating characteristic curve (AUROC) as the performance metric). We began with 10,000 random prediction pipelines which were evolved over 5 generations with an offspring size of 100 in each generation, using the recommended mutation rate of 0.9 and recombination rate of 0.1. The performance of each pipeline was evaluated using 20-fold stratified shuffled cross validation. This analysis revealed logistic regression with L2 regularization to be the most generalizable prediction model based on the cross-validation scores. To confirm the results of genetic programming, three additional methods including extreme gradient boosting implemented in XGBoost (Chen and Guestrin 2016), ensemble decision trees implemented in random forest (Svetnik, Liaw et al. 2003) and linear support vector machine (SVM) with linear kernel were also tested, using 20-fold stratified shuffled cross validation implemented in scikit-learn (Pedregosa, Varoquaux et al. 2011). The performance of machine learning models were evaluated using six metrics including AUROC, accuracy (number of correct predictions/total number of predictions), precision (True Positives / (True Positives + False Positives)), recall (True Positives / (True Positives + False Negatives)), F1 score (2*(Precision*Recall)/Precision+Recall), and balanced accuracy (bACC), the average of recall obtained on each class (i.e. severe/mild for baseline lung function and rapid/non-rapid for lung function decline).

To evaluate the statistical significance of the prediction performances (AUROC scores) obtained by ML models in comparison with random expectations, a non-parametric permutation test (Pedregosa, Varoquaux et al. 2011) was performed using 20-fold stratified shuffled cross-validation across hundred rounds of label switching and model training followed by empirical *p*-value estimation (i.e. the chance that the observed AUROC scores obtained using the data could be obtained by chance alone).

## Data Availability

All amplicon sequencing data generating in this project are deposited in NCBI GenBank under BioProject PRJNA763719.

## Acknowledgements

The project was supported by funding from CIHR (PJT-148827 to DN) and a Vertex Research Innovation Award (DN), and salary support from the Cystic Fibrosis Canada Research Fellowship (Award ID 558850 to JD), the Leopoldina Foundation (German National Academy of Sciences Leopoldina, Award ID LPDS 2017-17), the Reseau en Santé respiratoire (IL), and the Fonds de Recherche en Santé Quebec (IL, DN). MMS and BJS were supported by a Genome Canada and Genome Quebec Bioinformatics and Computational Biology grant. We would like to acknowledge Michael Surette for providing the PES genome sequence and Pradeep K. Singh for input in the Ampliseq design.

## Notes

### Competing Interest Statement

The authors have declared no competing interest.

### Author Declarations

The study was carried out with the approval from the Research Ethics Boards from the University of Calgary (15-0854) and McGill University Health Centre (15-623).

## Bibliography

Acosta, N., A. Heirali, R. Somayaji, M. G. Surette, M. L. Workentine, C. D. Sibley, H. R. Rabin and M. D. Parkins 2018. “Sputum microbiota is predictive of long-term clinical outcomes in young adults with cystic fibrosis.” Thorax 73(11): 1016–1025.

Acosta, N., F. J. Whelan, R. Somayaji, A. Poonja, M. G. Surette, H. R. Rabin and M. D. Parkins 2017. “The evolving cystic fibrosis microbiome: a comparative cohort study spanning 16 years.” Annals of the American Thoracic Society, 14(8): 1288–1297.

Alaa, A. M., & van der Schaar, M. 2018. Prognostication and risk factors for cystic fibrosis via automated machine learning. Scientific reports, 8(1), 1–19.

Aronesty, E. 2013. Fastq-mcf: sequence quality filter, clipping and processor.

Bouvier, N. M. 2016. Cystic fibrosis and the war for iron at the host–pathogen battlefront. Proceedings of the National Academy of Sciences, 113(6), 1480–1482.

Bragonzi, A., M. Paroni, A. Nonis, N. Cramer, S. Montanari, J. Rejman, C. Di Serio, G. Döring and B. Tümmler 2009. “Pseudomonas aeruginosa microevolution during cystic fibrosis lung infection establishes clones with adapted virulence.” American journal of respiratory and critical care medicine 180(2): 138–145.

Chen, T., & Guestrin, C. (2016, August). Xgboost: A scalable tree boosting system. In Proceedings of the 22nd acm sigkdd international conference on knowledge discovery and data mining (pp. 785–794).

Cingolani, P., A. Platts, L. L. Wang, M. Coon, T. Nguyen, L. Wang, S. J. Land, X. Lu and D. M. Ruden 2012. “A program for annotating and predicting the effects of single nucleotide polymorphisms, SnpEff: SNPs in the genome of Drosophila melanogaster strain w1118; iso-2; iso-3.” Fly 6(2): 80–92.

Coburn, B., P. W. Wang, J. D. Caballero, S. T. Clark, V. Brahma, S. Donaldson, Y. Zhang, A. Surendra, Y. Gong and D. E. Tullis 2015. “Lung microbiota across age and disease stage in cystic fibrosis.” Scientific reports 5(1): 1–12.

Cox, M. J., M. Allgaier, B. Taylor, M. S. Baek, Y. J. Huang, R. A. Daly, U. Karaoz, G. L. Andersen, R. Brown and K. E. Fujimura 2010. “Airway microbiota and pathogen abundance in age-stratified cystic fibrosis patients.” PloS one 5(6): e11044.

Cuthbertson, L., A. W. Walker, A. E. Oliver, G. B. Rogers, D. W. Rivett, T. H. Hampton, A. Ashare, J. S. Elborn, A. De Soyza and M. P. Carroll 2020. “Lung function and microbiota diversity in cystic fibrosis.” Microbiome 8: 1–13.

Dettman, J. R. and R. Kassen 2021. “Evolutionary genomics of niche-specific adaptation to the cystic fibrosis lung in Pseudomonas aeruginosa.” Molecular biology and evolution 38(2): 663–675.

Dias, R., & Torkamani, A. 2019. Artificial intelligence in clinical and genomic diagnostics. Genome medicine, 11(1), 1–12.

Emerson, J., Rosenfeld, M., McNamara, S., Ramsey, B., & Gibson, R. L. 2002. Pseudomonas aeruginosa and other predictors of mortality and morbidity in young children with cystic fibrosis. Pediatric pulmonology, 34(2), 91–100.

Eyre, D. W., D. De Silva, K. Cole, J. Peters, M. J. Cole, Y. H. Grad, W. Demczuk, I. Martin, M. R. Mulvey and D. W. Crook 2017. “WGS to predict antibiotic MICs for Neisseria gonorrhoeae.” Journal of Antimicrobial Chemotherapy 72(7): 1937–1947.

Firoz, A., M. Haris, K. Hussain, M. Raza, D. Verma, M. Bouchama, K. S. Namiq and S. Khan 2021. “Can Targeting Iron Help in Combating Chronic Pseudomonas Infection? A Systematic Review.” Cureus 13(3).

Flight, W. G., A. Smith, C. Paisey, J. R. Marchesi, M. J. Bull, P. J. Norville, K. J. Mutton, A. K. Webb, R. J. Bright-Thomas and A. M. Jones 2015. “Rapid detection of emerging pathogens and loss of microbial diversity associated with severe lung disease in cystic fibrosis.” Journal of clinical microbiology 53(7): 2022–2029.

Folkesson, A., Jelsbak, L., Yang, L., Johansen, H. K., Ciofu, O., Høiby, N., & Molin, S. 2012. Adaptation of Pseudomonas aeruginosa to the cystic fibrosis airway: an evolutionary perspective. Nature Reviews Microbiology, 10(12), 841–851.

Fothergill, J. L., Walshaw, M. J., & Winstanley, C. 2012. Transmissible strains of Pseudomonas aeruginosa in cystic fibrosis lung infections. European Respiratory Journal, 40(1), 227-238.

Goddard, A. F., B. J. Staudinger, S. E. Dowd, A. Joshi-Datar, R. D. Wolcott, M. L. Aitken, C. L. Fligner and P. K. Singh 2012. “Direct sampling of cystic fibrosis lungs indicates that DNA-based analyses of upper-airway specimens can misrepresent lung microbiota.” Proceedings of the National Academy of Sciences 109(34): 13769–13774.

Harris, J. K., B. D. Wagner, E. T. Zemanick, C. E. Robertson, M. J. Stevens, S. L. Heltshe, S. M. Rowe and S. D. Sagel 2020. “Changes in airway microbiome and inflammation with ivacaftor treatment in patients with cystic fibrosis and the G551D mutation.” Annals of the American Thoracic Society 17(2): 212–220.

Hisert, K. B., S. L. Heltshe, C. Pope, P. Jorth, X. Wu, R. M. Edwards, M. Radey, F. J. Accurso, D. J. Wolter and G. Cooke 2017. “Restoring cystic fibrosis transmembrane conductance regulator function reduces airway bacteria and inflammation in people with cystic fibrosis and chronic lung infections.” American journal of respiratory and critical care medicine 195(12): 1617–1628.

Javan, A. O., Shokouhi, S., & Sahraei, Z. 2015. A review on colistin nephrotoxicity. European journal of clinical pharmacology, 71(7), 801–810.

Jolly, A. L., D. Takawira, O. O. Oke, S. A. Whiteside, S. W. Chang, E. R. Wen, K. Quach, D. J. Evans and S. M. J. Fleiszig 2015. “Pseudomonas aeruginosa-induced bleb-niche formation in epithelial cells is independent of actinomyosin contraction and enhanced by loss of cystic fibrosis transmembrane-conductance regulator osmoregulatory function.” MBio 6(2).

Ke, Guolin, et al. “Lightgbm: A highly efficient gradient boosting decision tree.” Advances in neural information processing systems 30 (2017): 3146–3154.

Klockgether, J., N. Cramer, S. Fischer, L. Wiehlmann and B. Tümmler 2018. “Long-term microevolution of Pseudomonas aeruginosa differs between mildly and severely affected cystic fibrosis lungs.” American journal of respiratory cell and molecular biology 59(2): 246–256.

Koboldt, D. C., Q. Zhang, D. E. Larson, D. Shen, M. D. McLellan, L. Lin, C. A. Miller, E. R. Mardis, L. Ding and R. K. Wilson 2012. “VarScan 2: somatic mutation and copy number alteration discovery in cancer by exome sequencing.” Genome Research 22(3): 568–576.

Koehrsen, W. 2019. Feature Selector: Feature Selection in Python. https://github.com/WillKoehrsen/feature-selector

Kosorok, M. R., L. Zeng, S. E. West, M. J. Rock, M. L. Splaingard, A. Laxova, C. G. Green, J. Collins and P. M. J. P. p. Farrell 2001. “Acceleration of lung disease in children with cystic fibrosis after Pseudomonas aeruginosa acquisition.” Pediatric Pulmonology 32(4): 277–287.

Kresse, A. U., S. D. Dinesh, K. Larbig and U.J.M.m. Römling 2003. “Impact of large chromosomal inversions on the adaptation and evolution of Pseudomonas aeruginosa chronically colonizing cystic fibrosis lungs.” Molecular Microbiology 47(1): 145–158.

Kuhn, M. and K. Johnson 2013. Over-Fitting and Model Tuning. Applied Predictive Modeling. New York, NY, Springer New York: 61–92.

Kumru, B., Emiralioğlu, N., & Ozel, H. G. 2018. Does body mass index affect lung function in patients with cystic fibrosis?. Clinical Nutrition, 37, S91.

Le, T. T., W. Fu and J. H. Moore 2019. “Scaling tree-based automated machine learning to biomedical big data with a feature set selector.” Bioinformatics 36(1): 250–256.

Lees, J. A., Mai, T. T., Galardini, M., Wheeler, N. E., Horsfield, S. T., Parkhill, J., & Corander, J. 2020. Improved prediction of bacterial genotype-phenotype associations using interpretable pangenome-spanning regressions. Mbio, 11(4)

Li, H., B. Handsaker, A. Wysoker, T. Fennell, J. Ruan, N. Homer, G. Marth, G. Abecasis and R. Durbin 2009. “The Sequence Alignment/Map format and SAMtools.” Bioinformatics 25(16): 2078–2079.

Lim YW, Evangelista III JS, Schmieder R, Bailey B, Haynes M, et al. Clinical insights from metagenomic analysis of sputum samples from patients with cystic fibrosis. Journal of clinical microbiology 2014. 52:425–437.

Macesic, N., Don’t Walk, O. J. B., Pe’er, I., Tatonetti, N. P., Peleg, A. Y., & Uhlemann, A. C. 2020. Predicting phenotypic polymyxin resistance in Klebsiella pneumoniae through machine learning analysis of genomic data. Msystems, 5(3).

MacFadden, D. R., Melano, R. G., Coburn, B., Tijet, N., Hanage, W. P., & Daneman, N. 2019. Comparing patient risk factor-, sequence type-, and resistance locus identification-based approaches for predicting antibiotic resistance in Escherichia coli bloodstream infections. Journal of clinical microbiology, 57(6).

Mahé, P., & Tournoud, M. 2018. Predicting bacterial resistance from whole-genome sequences using k-mers and stability selection. BMC bioinformatics, 19(1), 1–11.

Marvig, R. L., H. K. Johansen, S. Molin and L. Jelsbak 2013. “Genome analysis of a transmissible lineage of Pseudomonas aeruginosa reveals pathoadaptive mutations and distinct evolutionary paths of hypermutators.” PloS genetics 9(9): e1003741.

Marvig, R. L., Sommer, L. M., Molin, S., & Johansen, H. K. 2015. Convergent evolution and adaptation of Pseudomonas aeruginosa within patients with cystic fibrosis. Nature Genetics, 47(1), 57.

Méric, G., L. Mageiros, J. Pensar, M. Laabei, K. Yahara, B. Pascoe, N. Kittiwan, P. Tadee, V. Post, S. Lamble, R. Bowden, J. E. Bray, M. Morgenstern, K. A. Jolley, M. C. J. Maiden, E. J. Feil, X. Didelot, M. Miragaia, H. de Lencastre, T. F. Moriarty, H. Rohde, R. Massey, D. Mack, J. Corander and S. K. Sheppard 2018. “Disease-associated genotypes of the commensal skin bacterium Staphylococcus epidermidis.” Nature Communications 9(1): 5034.

Mobegi, F. M., Cremers, A. J., De Jonge, M. I., Bentley, S. D., Van Hijum, S. A., & Zomer, A. 2017. Deciphering the distance to antibiotic resistance for the pneumococcus using genome sequencing data. Scientific reports, 7(1), 1–13.

Mowat, E., S. Paterson, J. L. Fothergill, E. A. Wright, M. J. Ledson, M. J. Walshaw, M. A. Brockhurst and C. Winstanley 2011. “Pseudomonas aeruginosa population diversity and turnover in cystic fibrosis chronic infections.” American journal of respiratory and critical care medicine 183(12): 1674–1679.

Pedregosa, F., G. Varoquaux, A. Gramfort, V. Michel, B. Thirion, O. Grisel, M. Blondel, P. Prettenhofer, R. Weiss and V. Dubourg 2011. “Scikit-learn: Machine learning in Python.” The Journal of machine Learning research 12: 2825–2830.

Pincus, N. B., E. A. Ozer, J. P. Allen, M. Nguyen, J. J. Davis, D. R. Winter, C.-H. Chuang, C.-H. Chiu, L. Zamorano and A. Oliver 2020. “A genome-based model to predict the virulence of Pseudomonas aeruginosa isolates.” Mbio 11(4).

Poudyal, B., & Sauer, K. 2018. The ABC of biofilm drug tolerance: the MerR-like regulator BrlR is an activator of ABC transport systems, with PA1874-77 contributing to the tolerance of Pseudomonas aeruginosa biofilms to tobramycin. Antimicrobial agents and chemotherapy, 62(2).

Raschka, S. 2018. MLxtend: Providing machine learning and data science utilities and extensions to Python’s scientific computing stack. Journal of open source software, 3(24), 638.

Recker, M., M. Laabei, M. S. Toleman, S. Reuter, R. B. Saunderson, B. Blane, M.E. Török, K. Ouadi, E. Stevens and M. Yokoyama 2017. “Clonal differences in Staphylococcus aureus bacteraemia-associated mortality.” Nature microbiology 2(10): 1381–1388.

Saber, M. M., & Shapiro, B. J. 2020. Benchmarking bacterial genome-wide association study methods using simulated genomes and phenotypes. Microbial genomics, 6(3).

Shanthikumar, S., M. N. Neeland, R. Saffery and S. Ranganathan 2019. “Gene modifiers of cystic fibrosis lung disease: a systematic review.” Pediatric pulmonology 54(9): 1356–1366.

Smith, E. E., D. G. Buckley, Z. Wu, C. Saenphimmachak, L. R. Hoffman, D. A. D’Argenio, S. I. Miller, B. W. Ramsey, D. P. Speert and S. M. Moskowitz 2006. “Genetic adaptation by Pseudomonas aeruginosa to the airways of cystic fibrosis patients.” Proceedings of the National Academy of Sciences 103(22): 8487–8492.

Snell, G. I., Bennetts, K., Bartolo, J., Levvey, B., Griffiths, A., Williams, T., & Rabinov, M. 1998. Body mass index as a predictor of survival in adults with cystic fibrosis referred for lung transplantation. The Journal of heart and lung transplantation: the official publication of the International Society for Heart Transplantation, 17(11), 1097–1103.

Somayaji, R., J. C. Lam, M. G. Surette, B. Waddell, H. R. Rabin, C. D. Sibley, S. Purighalla and M. D. Parkins 2017. “Long-term clinical outcomes of ‘Prairie Epidemic Strain’Pseudomonas aeruginosa infection in adults with cystic fibrosis.” Thorax 72(4): 333–339.

Stressmann, F. A., G. B. Rogers, C. J. Van Der Gast, P. Marsh, L. S. Vermeer, M. P. Carroll, L. Hoffman, T. W. V. Daniels, N. Patel and B. Forbes 2012. “Long-term cultivation-independent microbial diversity analysis demonstrates that bacterial communities infecting the adult cystic fibrosis lung show stability and resilience.” Thorax 67(10): 867–873.

Svetnik, V., Liaw, A., Tong, C., Culberson, J. C., Sheridan, R. P., & Feuston, B. P. 2003. Random forest: a classification and regression tool for compound classification and QSAR modeling. Journal of chemical information and computer sciences, 43(6), 1947–1958.

Turcios, N. L. 2020. Cystic fibrosis lung disease: An overview. Respiratory care, 65(2), 233–251.

Tümmler, B. 2006. Clonal variations in Pseudomonas aeruginosa. In Pseudomonas (pp. 35–68). Springer, Boston, MA.

Van Der Gast, C. J., A. W. Walker, F. A. Stressmann, G. B. Rogers, P. Scott, T. W. Daniels, M. P. Carroll, J. Parkhill and K. D. Bruce 2011. “Partitioning core and satellite taxa from within cystic fibrosis lung bacterial communities.” The ISME journal 5(5): 780–791.

Virtanen, P., R. Gommers, T. E. Oliphant, M. Haberland, T. Reddy, D. Cournapeau, E. Burovski, P. Peterson, W. Weckesser and J. Bright 2020. “SciPy 1.0: fundamental algorithms for scientific computing in Python.” Nature methods 17(3): 261–272.

Waskom, M. L. 2021. Seaborn: statistical data visualization. Journal of Open Source Software, 6(60), 3021.

Welsh MJ, Ramsey BW, Accurso F, Cutting GR. Cystic fibrosis. In: Scriver CR, Beaudet AL, Sly WS, Valle D, eds. Metabolic & molecular bases of inherited disease. 8th ed. Vol. 3. New York: McGraw-Hill, 2001:5121–5188.

Williams, D., B. Evans, S. Haldenby, M. J. Walshaw, M. A. Brockhurst, C. Winstanley and S. Paterson 2015. “Divergent, coexisting Pseudomonas aeruginosa lineages in chronic cystic fibrosis lung infections.” American journal of respiratory and critical care medicine 191(7): 775–785.

Zhang, L., & Mah, T. F. 2008. Involvement of a novel efflux system in biofilm-specific resistance to antibiotics. Journal of bacteriology, 190(13), 4447-4452.

Zhao, C. Y., Y. Hao, Y. Wang, J. J. Varga, A. A. Stecenko, J. B. Goldberg and S. P. Brown 2020. “Microbiome data enhances predictive models of lung function in people with cystic fibrosis.” The Journal of Infectious Diseases. 2020; jiaa655

Zhao, J., P. D. Schloss, L. M. Kalikin, L. A. Carmody, B. K. Foster, J. F. Petrosino, J. D. Cavalcoli, D. R. VanDevanter, S. Murray and J. Z. Li 2012. “Decade-long bacterial community dynamics in cystic fibrosis airways.” Proceedings of the National Academy of Sciences 109(15): 5809–5814.

