## Supplementary Figures and Tables for "Single nucleotide variants in *Pseudomonas aeruginosa* populations from sputum correlate with baseline lung function and predict disease progression in individuals with cystic fibrosis"

### Supplementary Tables and Figures

Supplementary Table 1. Definition of clinical and microbial factors evaluated for prediction of lung function severity and progression in CF patients.

| Clinical metadata | Definition |
|---|---|
| PFGE typing | Pulsed-Field-Gel Electrophoresis assessment of *Pa* genotype sa either Prairie Epidemic Strain (PES) or Unique. |
| Birth cohort | Patients grouped according to the date of birth. |
| Pseudomonas abundance | Relative abundance of *Pa* in sputum based on 16S rRNA sequencing (*Pseudomonas* vs total bacterial load reads). |
| Mucoid | Presence of mucoid colony morphology in dominant *Pa* strain (cultured from patient sputum). |
| Death | Recorded until end of data collection in 2017 (Deceased/ Not-deceased). |
| BMI | Body Mass Index. |
| Age | Age at time of sputum collection. |
| Host Genotype | CFTR genotype (ΔF508 homozygous/ΔF508 heterozygous and other types). |
| Biological sex at birth | Male or Female |
| Shannon | Shannon diversity index computed based on 16S rRNA amplicon sequencing data of the sputum microbiome. |
| Simpson | Simpson diversity index computed based on 16S rRNA amplicon sequencing data of the sputum microbiome. |

Supplementary Table 2. Performance metrics of different ML models for predicting baseline lung function (FEVp) and lung function decline.

| | | Genomic data (95% CI) | | | | Genomic and clinical data (95% CI) | | | |
|---|---|---|---|---|---|---|---|---|---|
| | | LR | SVC | RF | XGB | LR | SVC | RF | XGB |
| FEVp | AUROC | 0.87 (0.84,0.9) | 0.87 (0.84,0.89) | 0.76 (0.72,0.79) | 0.66 (0.62,0.7) | 0.92 (0.84,1) | 0.90 (0.85,0.99) | 0.83 (0.7,0.94) | 0.77 (0.62,0.88) |
| | bACC | 0.81 (0.78,0.84) | 0.79 (0.76,0.81) | 0.69 (0.66,0.73) | 0.61 (0.58,0.65) | 0.83 (0.72,0.94) | 0.84 (0.75,0.93) | 0.77 (0.67,0.87) | 0.7 (0.6,0.8) |
| | Accuracy | 0.81 (0.78,0.84) | 0.79 (0.76,0.81) | 0.69 (0.65,0.73) | 0.61 (0.57,0.65) | 0.83 (0.72,0.94) | 0.84 (0.75,0.93) | 0.76 (0.67,0.86) | 0.7 (0.6,0.8) |
| | F1 | 0.81 (0.78,0.83) | 0.78 (0.75,0.81) | 0.68 (0.65,0.72) | 0.6 (0.56,0.64) | 0.83 (0.72,0.94) | 0.83 (0.74,0.93) | 0.76 (0.66,0.86) | 0.69 (0.59,0.79) |
| | Precision | 0.83 (0.81,0.86) | 0.81 (0.79,0.84) | 0.71 (0.67,0.75) | 0.63 (0.59,0.68) | 0.84 (0.73,0.94) | 0.85 (0.76,0.93) | 0.8 (0.7,0.9) | 0.75 (0.64,0.87) |
| | Recall | 0.81 (0.78,0.84) | 0.79 (0.76,0.81) | 0.69 (0.65,0.73) | 0.61 (0.57,0.65) | 0.83 (0.72,0.94) | 0.84 (0.75,0.93) | 0.76 (0.67,0.86) | 0.7 (0.6,0.8) |
| Lung Decline | AUROC | 0.74 (0.71,0.78) | 0.68 (0.63,0.73) | 0.7 (0.66,0.75) | 0.62 (0.58,0.66) | 0.79 (0.7,0.88) | 0.69 (0.58,0.81) | 0.77 (0.63,0.85) | 0.66 (0.57,0.76) |
| | bACC | 0.63 (0.59,0.66) | 0.58 (0.55,0.61) | 0.58 (0.54,0.62) | 0.58 (0.55,0.61) | 0.66 (0.59,0.74) | 0.59 (0.56,0.62) | 0.63 (0.59,0.66) | 0.58 (0.54,0.62) |
| | Accuracy | 0.64 (0.6,0.67) | 0.61 (0.58,0.63) | 0.59 (0.56,0.63) | 0.6 (0.56,0.63) | 0.67 (0.6,0.75) | 0.61 (0.58,0.64) | 0.65 (0.61,0.68) | 0.6 (0.56,0.63) |
| | F1 | 0.62 (0.58,0.65) | 0.55 (0.52,0.59) | 0.57 (0.54,0.61) | 0.57 (0.54,0.61) | 0.66 (0.58,0.74) | 0.56 (0.52,0.6) | 0.62 (0.58,0.66) | 0.57 (0.54,0.61) |
| | Precision | 0.65 (0.61,0.69) | 0.59 (0.54,0.65) | 0.59 (0.55,0.63) | 0.6 (0.56,0.64) | 0.69 (0.6,0.78) | 0.59 (0.53,0.64) | 0.66 (0.62,0.71) | 0.61 (0.55,0.63) |
| | Recall | 0.64 (0.6,0.67) | 0.61 (0.58,0.63) | 0.59 (0.56,0.63) | 0.6 (0.56,0.63) | 0.67 (0.6,0.75) | 0.61 (0.58,0.64) | 0.65 (0.61,0.68) | 0.61 (0.56,0.63) |

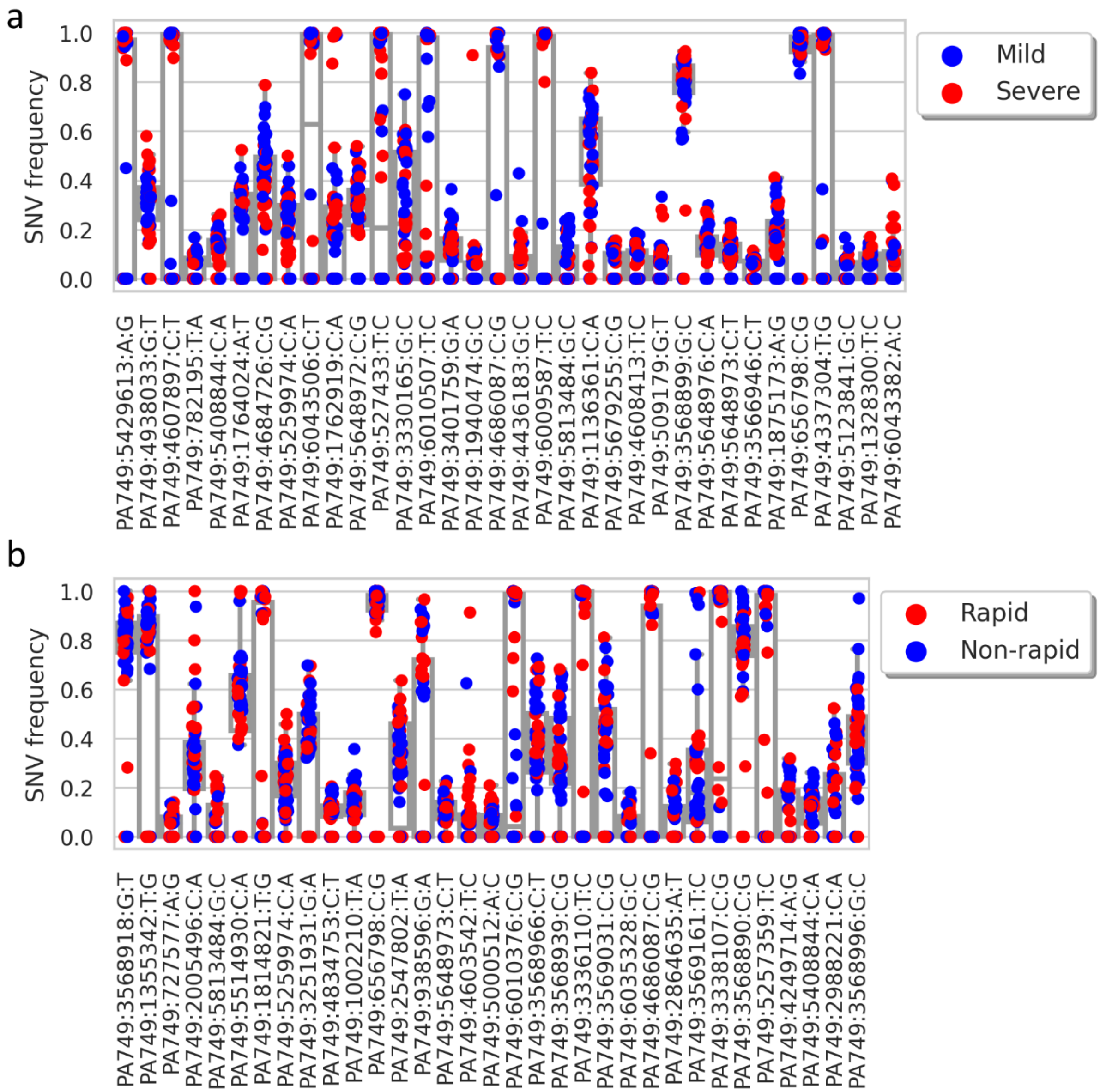


Supplementary Figure 1. Distribution of predictor SNV frequencies.

Frequency of identified predictor SNVs for (A) baseline lung function and (B) lung function decline across 54 patients. Samples are color-coded based on lung disease condition (Methods). Predictor SNVs are ordered based on their estimated feature importance (from high on the left, to low on the right) on the x-axis.

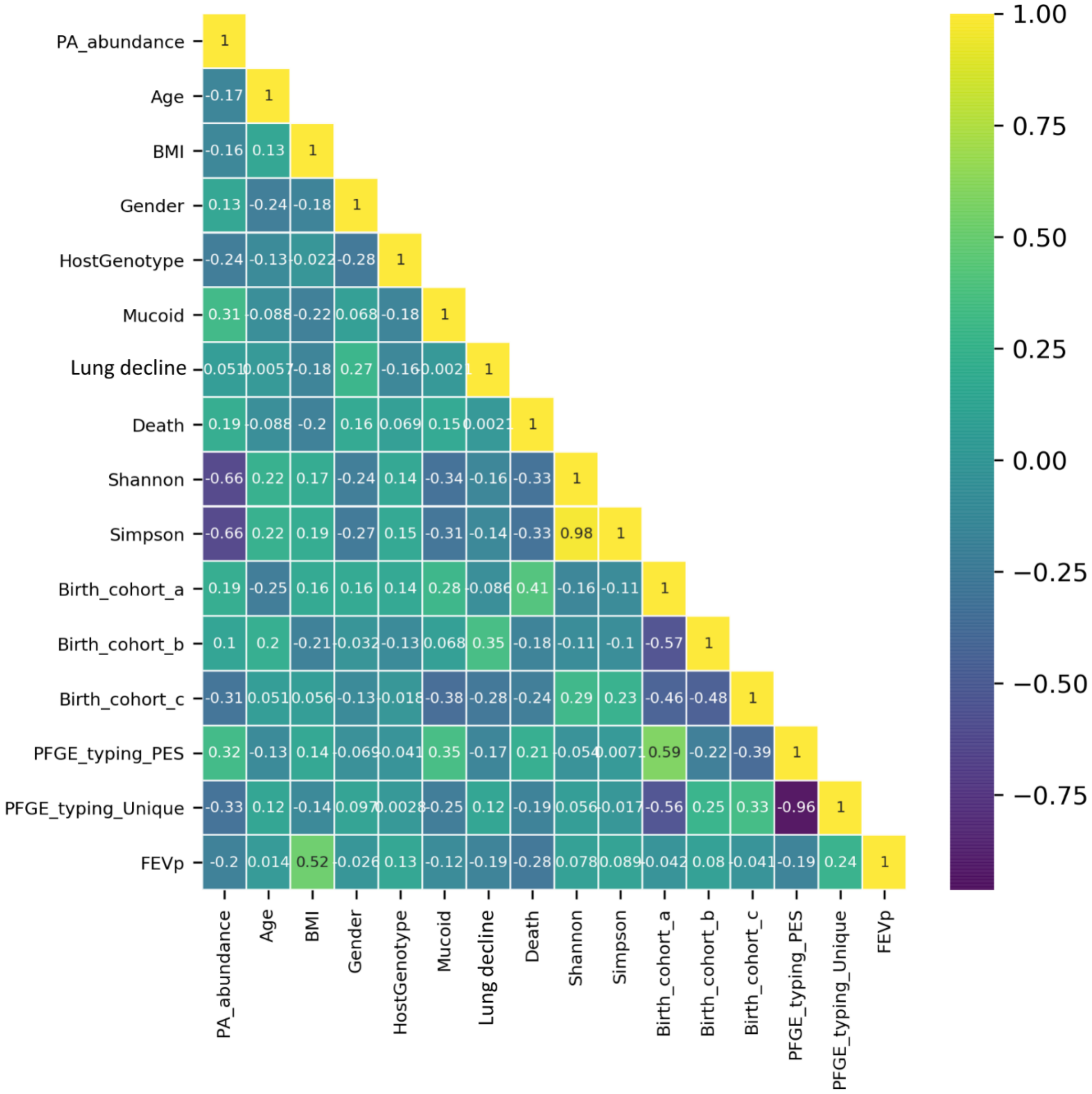


Supplementary Figure 2. Correlation between the evaluated clinical factors.

Pairwise Pearson correlation coefficients ($R^2$) heatmap of clinical data used prediction of lung disease severity and progression in CF patients.

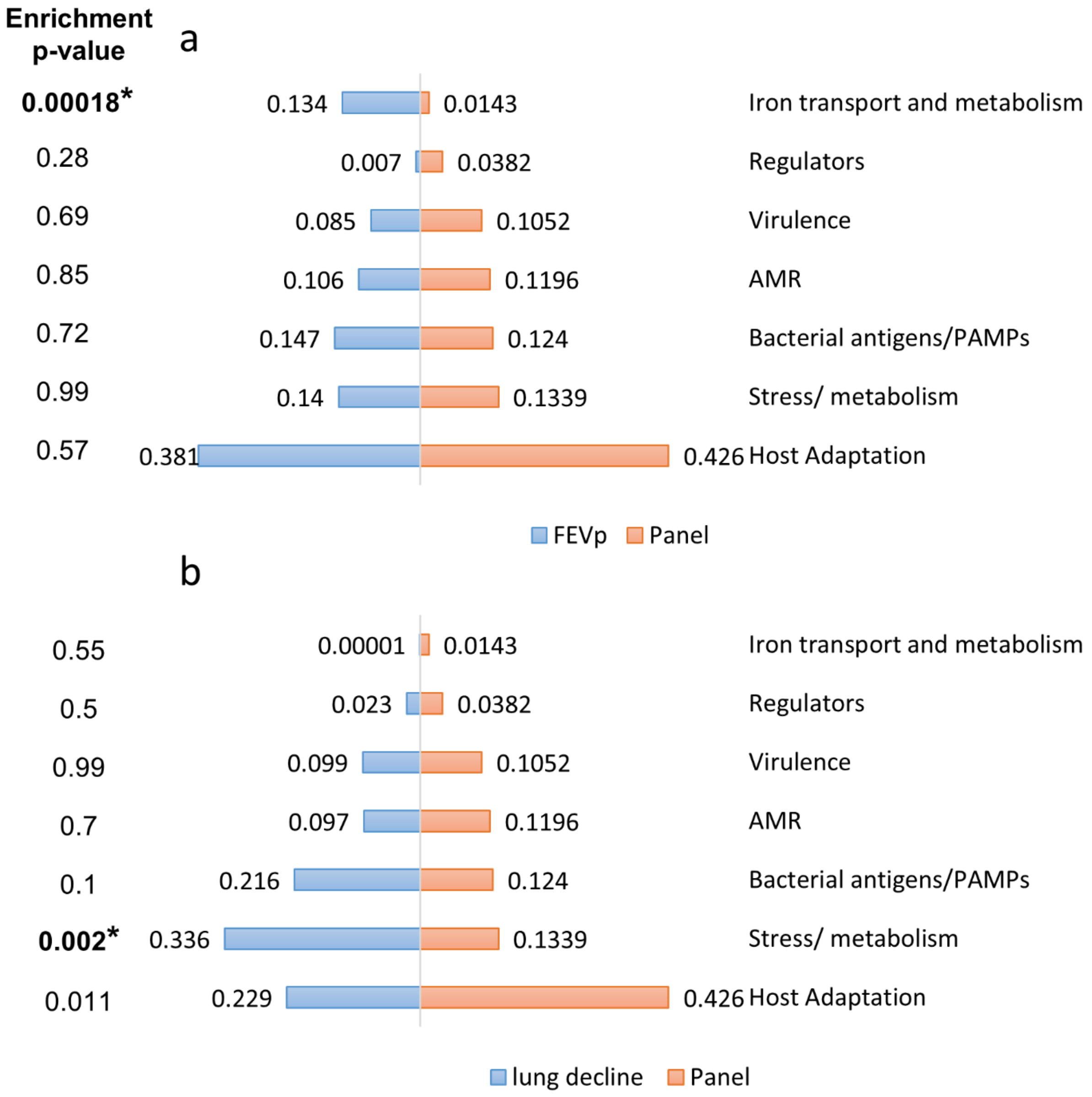


Supplementary Figure 3. Enrichment of predictor SNVs across functional categories in the AmpliSeq panel.

Predictor SNVs were functionally classified based on the function of genes harboring them (i.e. predictor genes) and enrichment of predictor genes for (a) baseline lung function (FEVp) and (b) progression (lung function decline) relative to total genes included in the AmpliSeq panel. Categories enriched in predictor genes relative to their expected frequency in the panel were identified using Fisher's Exact test. Statistically significant categories were determined at a significance level of $P < 0.05$ adjusted for multiple testing using the Bonferroni method (*i.e.* unadjusted $P < 0.005$).

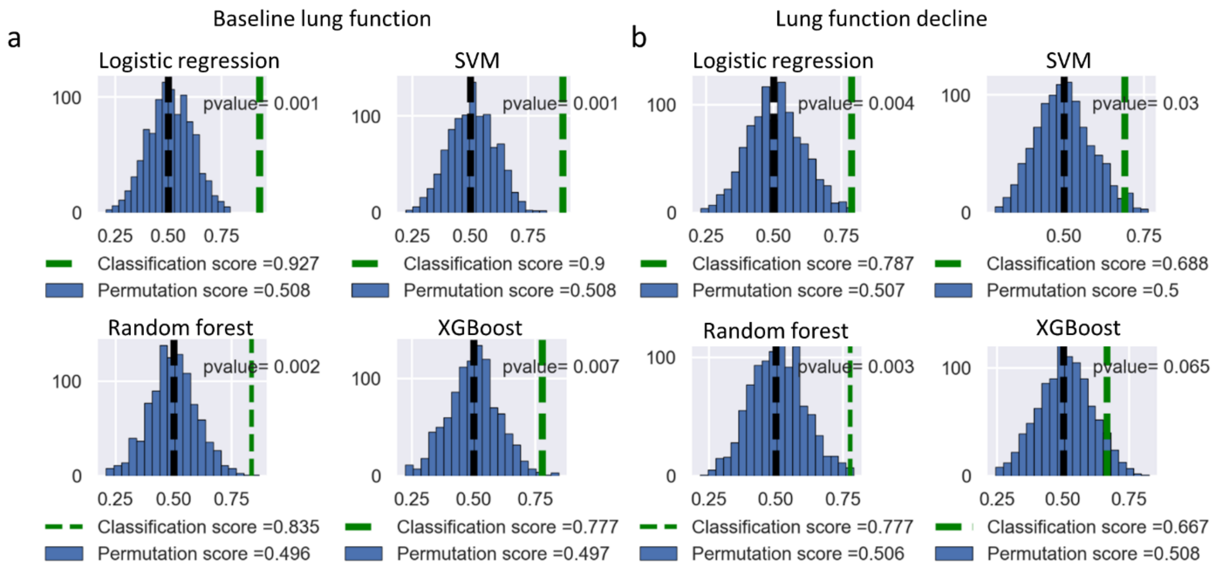


Supplementary Figure 4. Statistical significance of the performance of different ML models for prediction of lung disease severity and progression.

Average AUROC (area under the receiver operating characteristic curve) scores of four different ML models for (A) baseline lung function, and (B) lung function decline prediction compared with the same dataset with randomly shuffled labels.

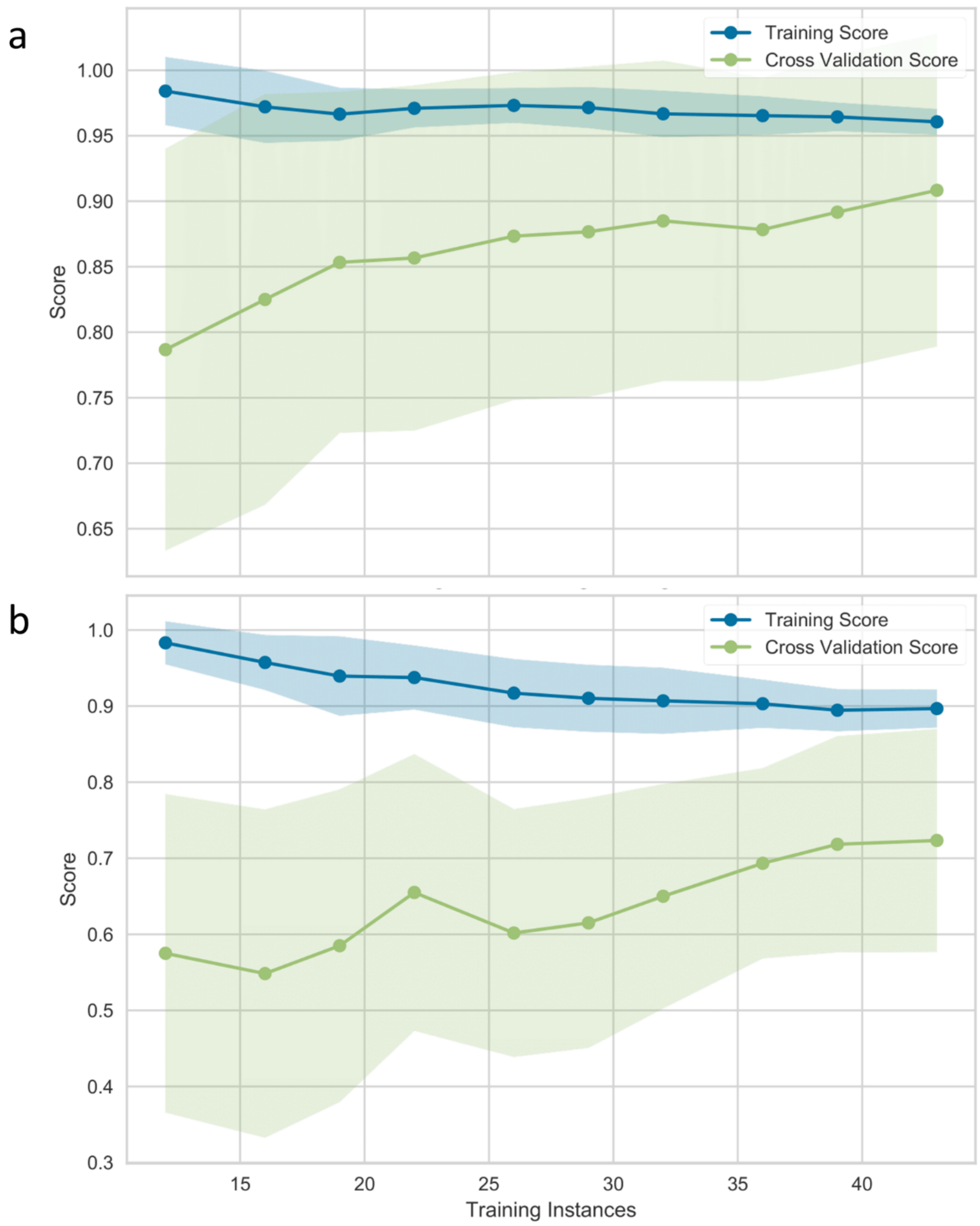


Supplementary Figure 5. Learning curve of logistic regression models for prediction of lung disease severity and progression.

Learning curves show the change in mean training accuracy (blue line) and cross-validation accuracy (green line) in predicting (A) baseline lung function, and (B) lung function decline as increasing numbers of samples are used to train the logistic regression model. Shading indicates the 95% confidence interval. Assessments at each number of training examples were through 20-fold stratified shuffled cross-validation.
